# Quantum Dot Encoding for In-Solution Single-Molecule Biomarker Counting in Metastatic Prostate Cancer

**DOI:** 10.1101/2025.11.03.25339402

**Authors:** Chia-Wei Kuo, Siva Nalla, Suresh Sarkar, Wonseok Lee, Liang Wang, Manish Kohli, Andrew M. Smith

## Abstract

Digital assays are in wide development for biomarker quantification at the single-molecule level, but the common use of surface-pulldown steps limits both analytical sensitivity and throughput. Here, we develop surface-free, wash-free, in-solution assays with a sensitivity slope approaching unity for sequence-specific counting of microRNAs (miRs) relevant to metastatic castration-resistant prostate cancer (mCRPC). These assays are enabled by DNA nanoflowers (DNFs) densely encoded with ∼200 fluorescent quantum dots (QDs) that assemble *in situ* stoichiometrically to miRs. The QD-DNFs are detected as single events in solution by fluorescence microscopy or flow cytometry without washing away unbound labels. A ∼50 aM limit of detection and high agreement with absolute target count (0.95) were achieved by machine learning-guided assay optimization, providing the potential for calibration-free measurements. Multiple miR sequences could be distinguished through ratiometric and colorimetric (5-color) QD signatures with a single excitation source for flexible detection scenarios in static solution or flow streams. The assays were applied for detecting exosomal miRs from small-volume plasma of mCRPC patients and showed strong agreement with RT-qPCR, but with more reliable detection of the trace prognostic biomarker miR-375. Consistent with our prior reports using large volume blood draws, higher plasma levels of miR-375 were associated with poor survival of patients with mCRPC. We anticipate that in-solution absolute counting of clinical biomarkers in plasma will enable robust molecular analysis of trace biomarkers needed for the translation of cancer precision medicine.

## INTRODUCTION

A new frontier of disease diagnosis and prediction is advancing based on low-abundance biomolecules from liquid biopsies, including circulating tumor DNA,^1–3^ tumor-associated microRNA (miR),^4–6^ and trace proteins like mutated p53.^7^ Traditional assays for these markers measure analog optical intensity of the labeled target,^8,9^ while newer single-molecule techniques are transitioning to digital detection and counting of individual molecular copies.^7,8,10,11^ Compared with analog assays, digital assays have an improved limit of detection (LOD)^7,8,11–13^ and potential for high-dimensional multiplexing due to the capacity to resolve single molecules.^13–15^ Most digital assay platforms are based on high-resolution microscopy after surface immobilization of the targets,^7,11,14,16,17^ which requires pull-down steps that are incomplete and variable, setting limits on sensitivity, while spatial scanning constrains throughput.^12,18,19^ In contrast, in-solution digital assays can comprehensively measure all biomarkers in a sample by diluting the target so that single copies are isolated within a single probe volume, analyzed either within microchambers, microdroplets, or a confocal volume of solution, or by counting in a flow stream.^1,4,20,21^

A key hurdle to in-solution assays is the absence of “wash” steps to remove unbound optical labels to reveal target molecules that are specifically labeled.^12,25^ The most advanced format is droplet digital polymerase chain reaction (ddPCR) for nucleic acid counting in microdroplets,^22,23^ which achieves molecular counting by a combination of two signal amplification processes. One component is a “turn-on” fluorescent sensor dye that increases in intensity by an order of magnitude upon binding to targets,^22,24^ which is applied together with *in situ* exponential production of additional targets by PCR. However, the low availability of spectrally distinguishable sensor dyes sets the limit of multiplexing of ddPCR^4,15,21^ although advances continue to be pursued.^23,24^ Alternatively, broad classes of spectrally tunable dyes can be applied for multiplexed in-solution “wash-free” digital assays by binding multiple labels to each target,^1,20,26^ which then registers as a brighter molecular entity that can be measured by methods like fluorescence correlation spectroscopy (FCS).^27,28^ However, unbound labels set the LOD typically to the picomolar range, at least three orders of magnitude above the range of trace biomarkers in serum,^4,20,25^ and large solution volumes must be analyzed, which is time-consuming. In such scenarios that may allow multiplexing, targets must assemble labels in great excess relative to the ambient solution so that off-target counts are near zero to achieve a LOD appropriate for trace analytes.

Here, we introduce a self-assembly and readout process called iQ-Flow which generates fluorescently labeled nucleic acid targets detectable far above background levels of unbound labels without washing, with rapid readout in a flow stream. We focus on miR biomarkers, a class of short (∼22 nucleotides) non-coding RNAs that we previously showed predict progression of patients with metastatic castration-resistant prostate cancer (mCRPC).^33–35^ The miRs are stoichiometrically grown into DNA nanoflowers (DNFs) that are larger than the optical diffraction limit (∼200 nm) for single-molecule imaging^29^ and particle size thresholds (500–1000 nm) for flow-counting using a benchtop flow cytometer.^30^ We use nanometer-scale fluorescent quantum dots (QDs) to optically encode the DNFs, targeting attachment of >100 QDs for precise optical codes. To measure multiple targets simultaneously, multicolor QDs self-assemble sequence-specifically with DNFs to generate multispectral codes with single-wavelength excitation.^31^ Because labeling is limited by QD steric hindrance,^32^ we develop means to disassemble and reassemble the QD-DNFs to enhance labeling density. We apply iQ-Flow to extracts of small-volume blood plasma, targeting miR biomarkers present at low concentrations (attomolar to femtomolar) with high inter-sample variability.^36,37^ We benchmark the study in comparison with gold-standard reverse transcriptase quantitative PCR (RT-qPCR) and analyze predictive metrics in a small cohort study of mCRPC patients.

## RESULTS AND DISCUSSION

### iQ-Flow Assay Design

The goal is to extend miRs into long, single-stranded DNA (ssDNA) composed of hundreds of repeating sequences so that they can be labeled by a large number of QDs conjugated to complementary sequences. As shown in **Figure 1a**, a target miR is first sequence-specifically extended through rolling circle amplification (RCA) using a circular template generated by ligation of a 121-base linear padlock DNA probe (**Figure S1** and **Table S1**). By controlling the RCA time, a DNA nanoflower (DNF) grows from each target miR as a folded, quasi-spherical product with tunable size in the range of hundreds of nanometers, composed of 121-base repeats. QDs with ∼10 nm sizes are conjugated to complementary sequences that hybridize with the DNF, yielding a fluorescent spot readily detected in solution *via* hyperspectral confocal microscopy (**Figure 1b**) or flow cytometry (**Figure 1c**). When detected in a flow stream, we call this full process iQ-Flow.

**Figure 1.**
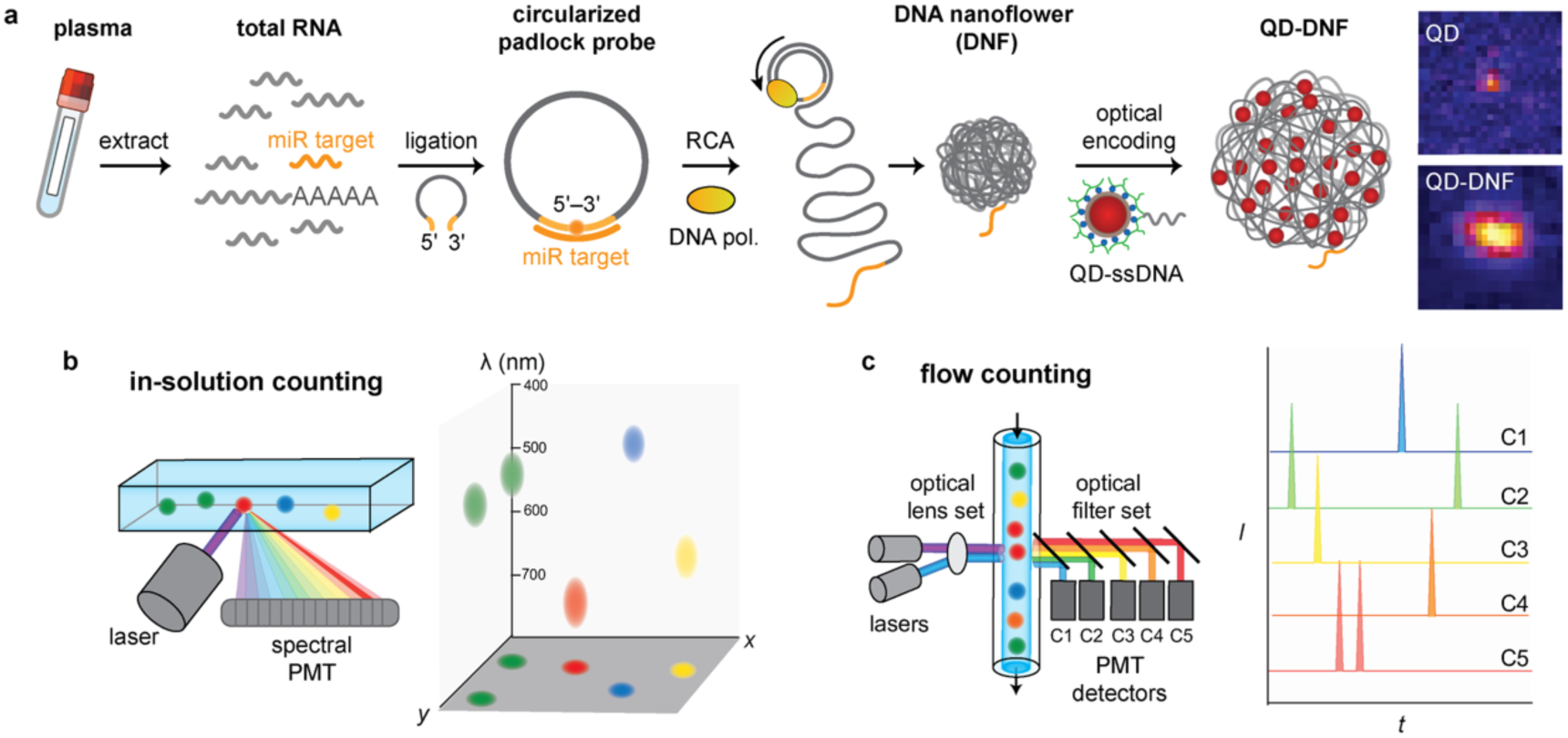
iQ-Flow for single-molecule in-solution nucleic acid quantification. **(a)** Assay workflow showing RNA extraction from blood plasma and extension of one miR target sequence through circularization of an ssDNA padlock probe for rolling circle amplification (RCA). The product is a submicron DNA nanoflower (DNF) containing repeating sequences complementary to the template. The DNF is then labeled in a sequence-specific manner with quantum dot (QD)-ssDNA conjugates. Sequences are in **Table S1**. Fluorescence micrographs show a large, bright spot for QD-DNF compared with QD alone. **(b)** Schematic of in-solution counting by microscopy with a single excitation source and a multispectral photomultiplier (PMT) detector, yielding a multispectral image of single QD-DNFs. **(c)** Schematic of in-solution flow-counting with a single excitation source and fluorescence detectors (channels C1 to C5), yielding rapid time-traces of QD-DNFs.

### On-Surface *vs.* In-Solution Assays

To test if there are intrinsic advantages to in-solution or on-surface readouts of QD-DNFs, analytical sensitivities were compared in the two test formats. QD-DNFs were imaged either in solution by confocal microscopy (**Figure 2a**) or after surface capture by total internal reflectance fluorescence (TIRF) microscopy (**Figure 2b**). Fluorescent spots with sizes near 400 nm (**Figure 2c-d**) were counted using a spot identification algorithm with empirically optimized detection thresholds (**Figure S2, S3**).^38^ An assay with perfect analytical sensitivity has a 1:1 relationship between spot count and dilution factor with a slope (*m*) of 1.0. The in-solution assay, including excess free labels in solution, had a near-ideal slope (*m* = 1.2) and agreement with ideal readout (intraclass correlation coefficient, ICC = 0.95) over a ∼10^3^ linear dynamic range (**Figure 2e**) with absolute counts close to the theoretical number of DNFs in the solution volume. For the surface-based assay, all values were lower (*m* = 0.46, ICC = 0.65, **Figure 2f**), even with unbound labels washed away, and absolute counts were hundreds to thousands of times lower than the DNF number in solution. A diminished slope is commonly observed when pull-down is applied^7,8,12,39,40^ and is attributed to cumulative repulsion between captured targets as well as the depletion of capturing agents (**Figure S4**)^41^ and limits the capacity to distinguish differences in analyte concentration.

**Figure 2.**
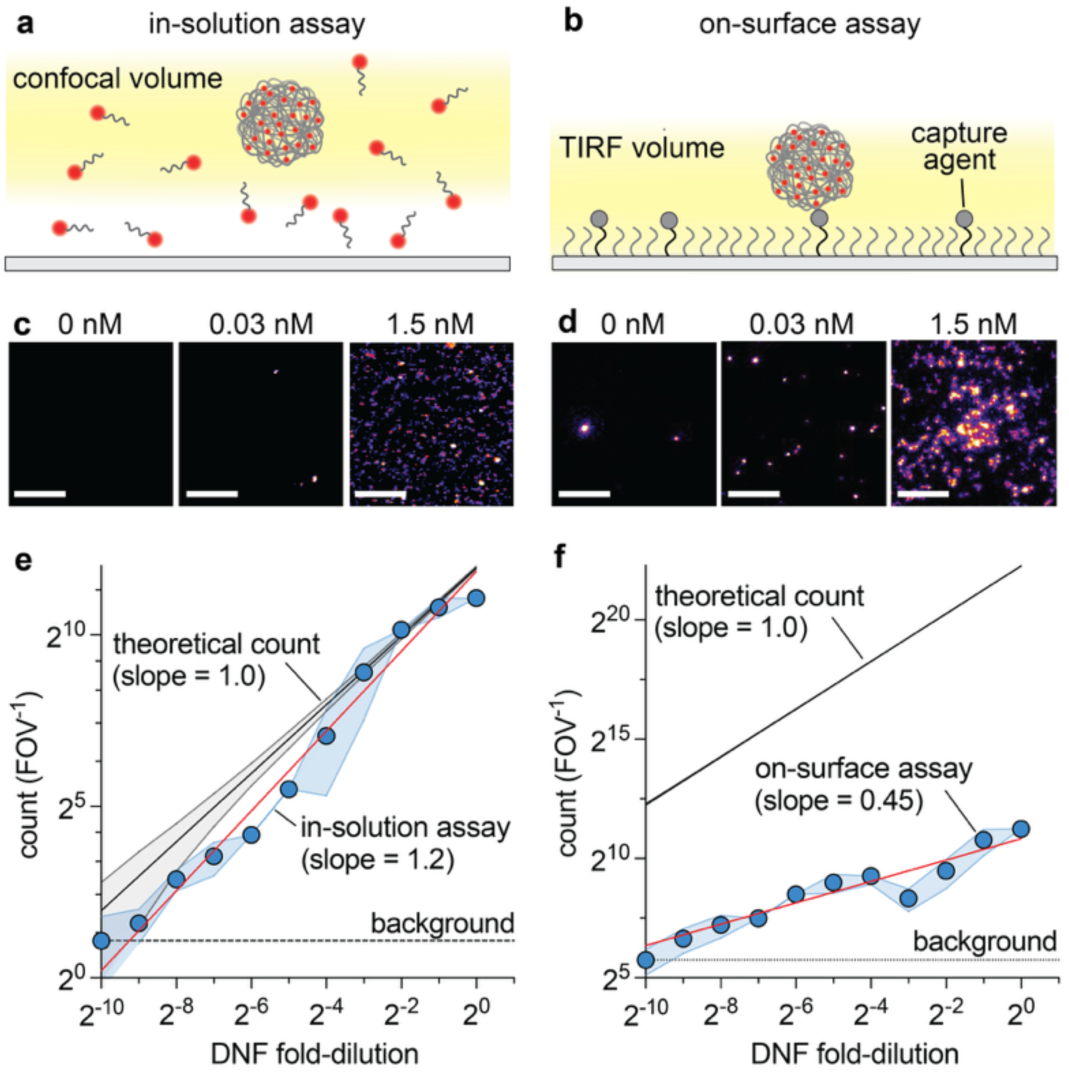
Analytical sensitivity is higher for in-solution digital counting of labeled DNFs compared with counting after surface capture. **(a)** Labeled DNFs imaged in solution (glycerol-water mixture) by confocal fluorescence microscopy with excess unbound labels remaining in solution. **(b)** Labeled DNFs captured on a surface and imaged through TIRF microscopy with unbound labels washed away. **(c,d)** Representative fluorescence micrographs for the two measurements at the indicated DNF concentration. Scale bar: 5 µm. **(e,f)** Digital counts per field of view (FOV) *versus* DNF dilution factor (1.5 nM solution). Data points are mean counts per FOV with standard deviation (S.D.) shown as shading, applying optimized spot detection thresholds (15 dB for on-surface, 3 dB for in-solution; **Figure S3**). *N* = 9. Red lines are linear regressions of log-log-transformed data with the indicated slope. Black lines are theoretical counts based on the DNF number in the solution, with grey shading showing confidence intervals (C.I.) The black dashed line indicates the background count.

### QDs Enhance In-Solution Digital Detection

DNF detection in the presence of unbound fluorescent labels was dependent on the nature of the fluorophore. Fluorescent dyes covalently attached to ssDNA complementary to the DNF were compared with wavelength-matched QDs comprising ∼5 nm core/shell CdSe/CdZnS bound to ssDNA through click chemistry to a multidentate polymer adsorbed to the nanocrystal surface (**Figure S5, S6**).^42^ With both organic dyes and semiconductor QDs, labeled DNFs were readily detected above background signals in solution by confocal microscopy, even when outnumbered 200:1 by unbound labels (**Figure 3a-d**). QDs provided a higher mean brightness (12.6 ± 2.0 a.u.) compared with dyes (8.69 ± 0.94 a.u.) and a lower dependence on the spot detection threshold (*m* = –0.010) compared with dyes (*m* = –0.015), indicating more robust detection (**Figure 3e**). This is important because single-molecule assays require the application of an arbitrarily chosen detection threshold to balance sensitivity and specificity, and a smaller dependence is preferred. This outcome may derive from a reduced propensity of QDs to self-quench compared with dyes when densely labeling a single molecule.

**Figure 3.**
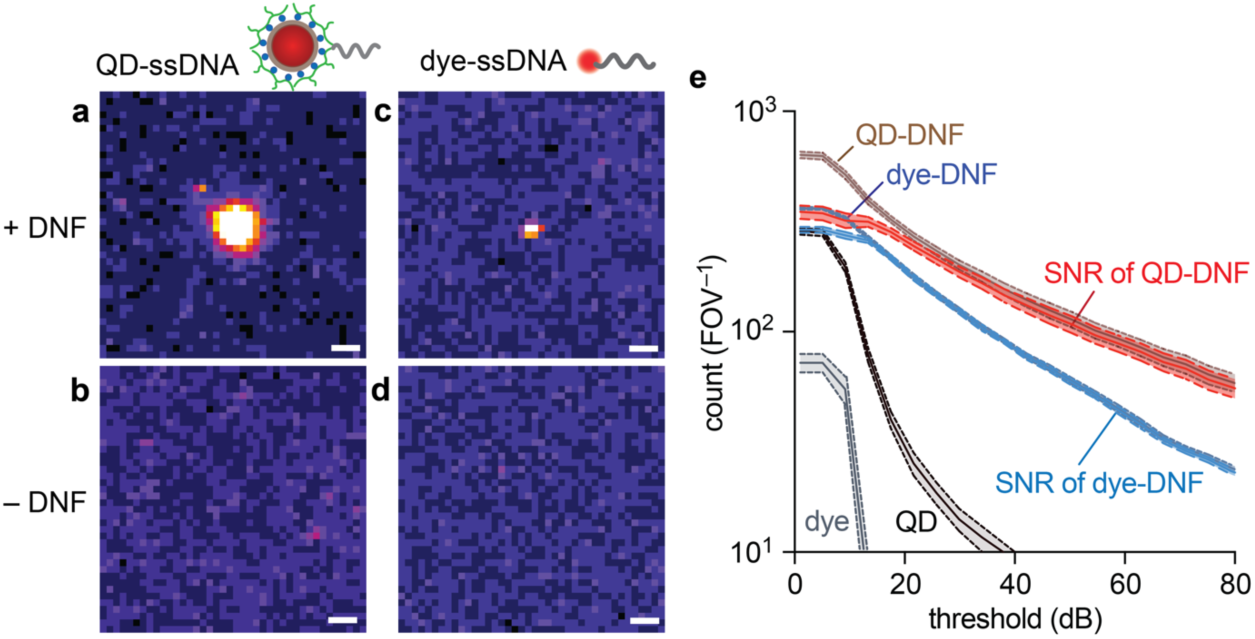
DNF counting in-solution comparing dye- or QD-based fluorescent labels. **(a-d)** Representative confocal fluorescence micrographs of QD-ssDNA and dye-ssDNA and without DNFs present in glycerol-water solutions. Both fluorophores emit at 605 nm. Scale bar: 1 µm. Uncropped fields of view are shown in **Figure S7**. **(e)** Spot counts for DNFs labeled with dye-ssDNA (blue) or dye-ssDNA alone (grey), as well as DNFs labeled with QD-ssDNA (brown) or QD-ssDNA alone (black) for the indicated spot detection thresholds. The signal-to-noise ratios (SNR) are shown for dyes (light blue) and QDs (red) as the difference between counts with and without DNFs. Shading indicates the S.D. of counts between different fields of view. *N* = 9.

### Steric Hindrance Limits QD Labeling Density

Maximizing the number of labels per DNF is necessary for high-sensitivity counting amid a background of free labels. We found that through hybridization with DNFs, dyes with ∼1 nm size consistently out-labeled ∼13 nm QDs by a factor of ∼2, with QDs plateauing at a maximum mean near 58 (**Figure 4a, 4b**, and **S8**), with log-normal distributions (**Figure S9**). When the DNF was first labeled with ssDNA spacers^44^ designed to separate the DNF and QD by ∼20 nm there was a 1.4-fold increase in QDs per DNF (**Figure 4c** and **S10**), although many DNFs remained poorly labeled (52 ± 9 % with <50 QDs). The low labeling density therefore may be driven by steric hindrance, which is consistent with a negative monotonic trend between QDs per DNF and hydrodynamic diameter (**Figure 4d-f**).^42,43^ The smallest size (10.4 nm) yielded a similar number of labels per DNF as the dye (Student’s *t*-test; *p* > 0.05), while half-maximal label density was for QDs near 13.7 nm, similar to that previously observed for QD labeling of mRNA in cells, potentially reflecting a pore size in long, single-stranded nucleic acids.^32^

**Figure 4.**
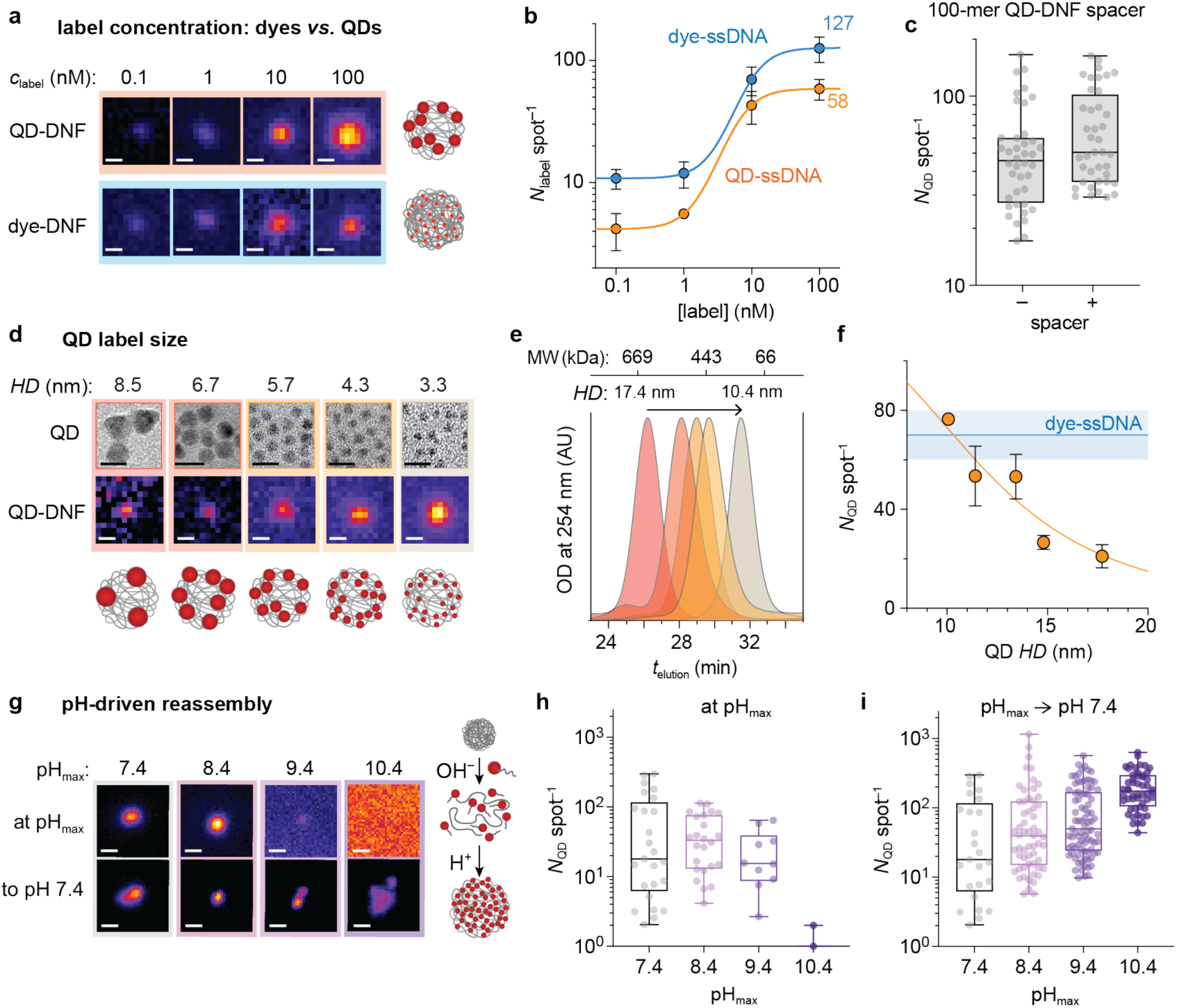
Optimizing QD labeling of DNFs to overcome steric hindrance. **(a)** Representative fluorescence micrographs show DNFs labeled with dye-ssDNA (blue) or QD-ssDNA (orange) at the indicated label concentrations. Scale bar: 0.5 µm. **(b)** Labels per spot at indicated concentration (mean ± S.D., *N* = 3). **(c)** QDs per DNF with and without 100-mer ssDNA spacers. Boxes indicate 25/75 percentiles with the enclosed line indicating the mean. Whiskers are the maximum and minimum values. *N* = 3. **(d)** Transmission electron microscope (TEM) micrographs and mean diameters for a size series of QDs, and representative fluorescent micrographs showing the impact of QD size on labeling density. Schematics showing the impact of QD size on labeling density. Scale bar: 10 nm (TEM) and 0.5 µm (fluorescence micrograph). **(e)** Gel permeation chromatograms for polymer-coated QDs showing hydrodynamic diameter (*HD*) of 10.4 to 17.4 nm for QDs in panel (d), with elution times of globular protein standards shown on the top *x*-axis. OD = optical density. AU = arbitrary units. **(f)** Number of QDs per DNF spot *versus* QD *HD* with logistic regression (orange line). Blue line indicates dyes per spot for equivalent experimental conditions (mean ± S.D., *N* = 3). **(g)** pH-driven reassembly of DNFs from pH 7.4 to pH_max_, and back to pH 7.4, showing representative fluorescence micrographs of QD-DNFs. Scale bar: 0.5 µm. The schematic shows that the DNF expands to allow QDs greater labeling access at elevated pH prior to restoring to pH 7.4 to allow hybridization. **(h)** Number of QD per spot at the indicated pH_max_. **(i)** Number of QD per spot at pH 7.4 after treatment at the indicated pH_max_. The boxes show the 25/75 percentiles with the mean as the enclosed line. Whiskers are the maximum and minimum values.

### Maximizing QD-DNF Labeling

Although the smallest QDs resulted in DNF labeling similar to dyes, their brightness levels are low and their spectral bands are less tunable and broader than those of larger QDs (**Figure S11**), limiting multiplexing capacity. Therefore, we aimed to overcome steric hindrance for larger QDs (*HD* = 13.4 nm) which have highly tunable and homogeneous spectral features. Assuming that QDs cannot access the DNF interior due to its condensed structure,^45^ we devised a reversible DNF denaturation process by increasing the pH (**Figure 4g-i**) from 7.4 to an alkaline value of pH_max,_ to expand its ssDNA coils.^46^ When pH_max_ reached 10.4, fluorescent spots were no longer detectable amid a bright fluorescent haze, but spots reappeared when the pH was returned to 7.4 (**Figure 4g**).

Applying this process, the mean labeling density rose 16-fold, from ∼9 QD spot^−1^ to 149 QD spot^−1^ (**Figure S12**), and intensity distributions became uniform (coefficient of variation, C.V. = 13.2%) compared to single-pH labeling (C.V. = 50.4%). Most importantly, the number of spots with low labeling density decreased substantially with increasing pH_max_, with none detected below 23 QD spot^−1^.

### Structure of QD-DNF Nanocomposites

The DNF structure was evaluated before and after labeling with QD-ssDNA. DNFs can be tuned across a wide range of sizes from tens of nanometers to microns by the duration of RCA.^47^ A 4-hour reaction resulted in the highest brightness (229.5 QD spot^−1^) while maintaining a uniform intensity distribution (C.V. = 13.7%; **Figure S13**). Scanning electron microscopy (SEM) showed DNFs to be near 500 nm (**Figure 5a**) after drying on a substrate, with flower-like structures and apparent surface pores. After pH-reassembly-based QD-ssDNA labeling, QD-DNFs occupied a 5-fold larger area with less porous surface decorated with protrusive features near 10 nm, matching the sizes of QDs (**Figure 5b, 5c**). These structures in the dry state differed from those measured in solution by FCS, with a DNF *HD* of 106 ± 28 nm measured when labeled with intercalating dyes and QD-DNF near 1330 ± 110 nm (**Figure 5d**).^48,49^ QD-DNFs were much larger than DNFs labeled with dye-ssDNA (146 ± 38 nm), likely because QDs are more than 1000-fold larger in volume than dyes (**Figure 5d** and **S14**). These in-solution FCS sizes were consistent with spot sizes by fluorescence microscopy (**Figure 4g**) and, importantly, compatible with micron-scale detection limits of benchtop flow cytometers.

**Figure 5.**
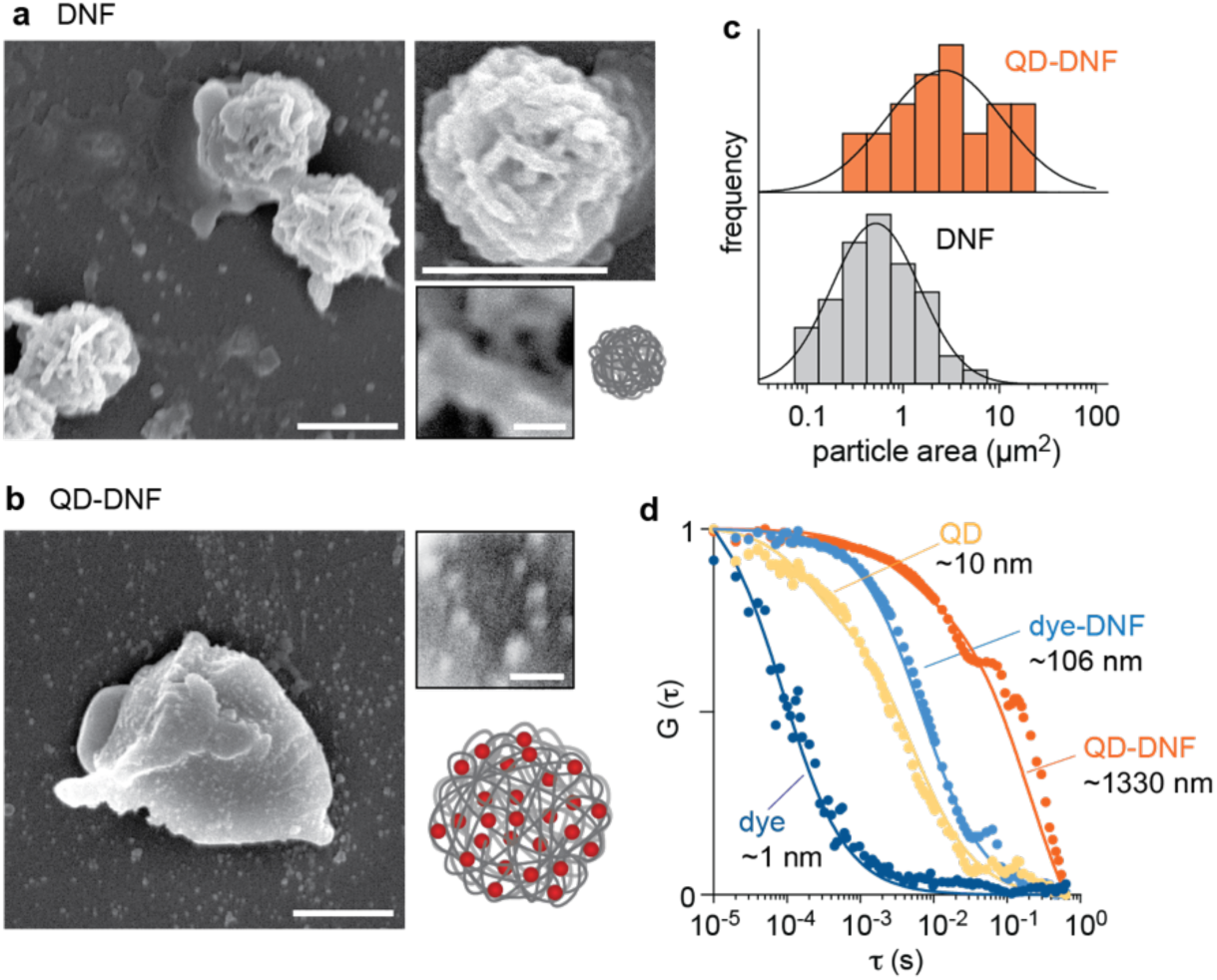
QD-DNF nanocomposite structure. Representative scanning electron micrographs show **(a)** DNF and **(b)** QD-DNF with zoomed-in regions showing expanded views of the particle surfaces. Scale bar: 500 nm (main panel) and 20 nm (inset). **(c)** SEM particle area distributions for DNFs and QD-DNFs with black lines showing lognormal fits. Data were collected from 10 images. **(d)** Representative FCS autocorrelation curves and calculated hydrodynamic diameter for intercalating dye, QD-ssDNA, and DNF labeled with intercalating dye or QD-ssDNA, each in aqueous solution.

### Assembly of Multicolor QD-DNFs for Colorimetric Multiplexing

We designed a scheme for in-solution miR counting of multiple distinct miR sequences in the same mixture using miR-templated DNFs that self-assemble sequence-specifically with different colors of QD-ssDNAs (**Figure 6a** and **Table S1**). Five QDs (QD445, QD525, QD570, QD605, and QD625) were synthesized with spectrally distinct emission bands (**Figure 6b** and **Figure S15a**)^43,50,51^ by tuning both composition and size (**Figure S15b**), and further conjugated to distinct ssDNA sequences (R1’ to R5’; **Table S1**). DNFs with 5 repeated labeling sequences (L1’ to L5’; **Table S1**) specifically assembled with color-specificity in a mixture of the 5 QD-ssDNA labels and their corresponding spacers (L1–R1 to L5–R5), yielding bright, spectrally distinct spots amid an excess of multicolor labels (**Figure 6c-d**). Due to the large number of QDs assembled, even QD emission bands with 47% overlap (QD605 and QD625) at the ensemble level were distinguishable (**Figure 6e**) with <5 % false positive count rates (**Figure 6f**). With ∼200 QDs per QD-DNF, multiple distinct QD colors could also precisely assemble into a single DNF to provide ratiometric codes to expand multiplexing capacity (**Figure 6g**). For multicolor labeling, when QDs were different in size (QD570 with QD625; **Figure S15b**), achieving a matching intensity required pH-driven reassembly (**Figure 6h-i**), as the larger QD625 was selectively excluded from the DNF (**Figure S16**).

**Figure 6.**
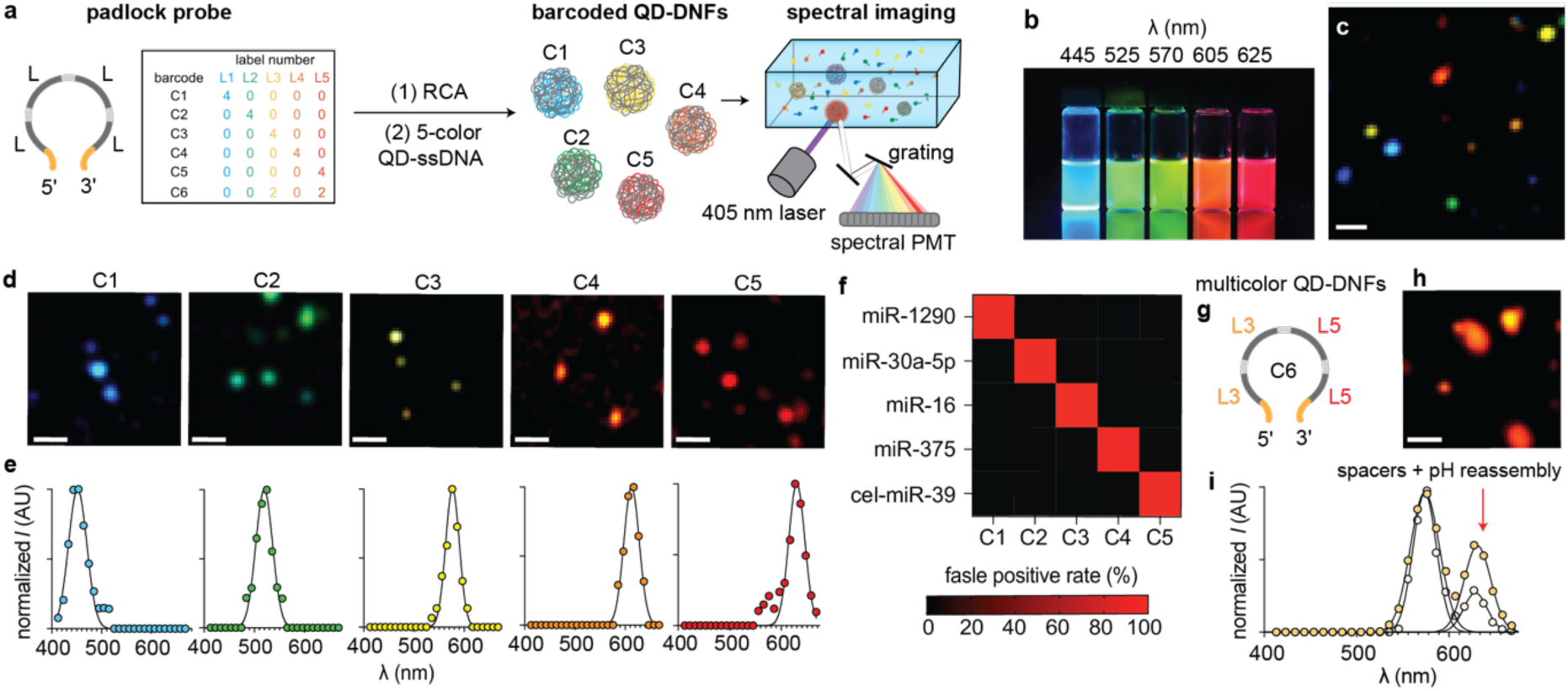
Multiplexed single-molecule counting in-solution by colorimetric and ratiometric labeling of DNFs. **(a)** Schematic shows ssDNA template with 4 labeling regions (gray color) and miR target region (yellow) used for miR-dependent elongation by RCA. Resulting DNFs are labeled with a 5-color mixture of QDs conjugated to complementary ssDNA. Colorimetric barcodes are determined in solution using a single excitation source (405 nm laser) and a hyperspectral photomultiplier tube (PMT) array. **(b)** Solutions of colloidal QDs showing fluorescence under ultraviolet excitation. The number indicates the peak emission wavelength of each QD sample. **(c)** A representative hyperspectral micrograph shows a mixture of spectrally barcoded QD-DNFs in solution. **(d)** Representative multispectral micrographs using a mixture of the 5 QDs shown in panel (b) with only a single DNF target added in each case. **(e)** Representative fluorescence spectra of detected spots from panel (d). A Gaussian fit (black) is shown for each spectrum. **(f)** Confusion matrix for in-solution quantification of 5 different miRs (miR-1290, miR-30a-5p, miR-16, miR-375, and cel-miR-39) measured in each of 5 color-code channels. The false positive rate of detection is indicated in the color scale. *N*=5. **(g)** Schematic shows the padlock probe template for ratiometric barcoding for an equal number of labeling regions L3 and L5. DNF products were labeled with a mixture of QD570– R3’, QD625–R5’, and their corresponding spacers (L3–R3 and L5–R5). **(h)** Representative multispectral micrograph of ratiometrically barcoded QD-DNFs. **(i)** Representative fluorescence spectra of detected spots from panel (h), comparing single-pH labeling without spacers *versus* pH-driven reassembly with spacers. Additional fluorescence micrographs are in **Figure S16**. Scale bars = 5 µm. AU = arbitrary units.

### iQ-Flow with Flowstream Readout

We optimized iQ-Flow for single-molecule counting at high-throughput using a benchtop flow cytometer (**Figure 7a**).^4^ For high-fidelity counting across a wide concentration range, we applied two spectrally distinct QD labels for each target (QD605, QD700) together with an intercalating dye (SYBR Green; **Figure 7b**). A high degree of colocalization was observed between each spectral channel and with side scattering (**Figure 7c**). A supervised machine learning process (described in **Supporting Methods**, **Figures S17-S19**) was used to optimize the labeling reagents to achieve a LOD of 47 aM (**Figure 7d**), a 10^3^-fold improvement over initial designs and around 10^5^-fold better than the LOD of RCA by ensemble fluorescence (**Figure S20**). The dynamic range spanned more than 5 orders of magnitude, with differentiation between certain miR isoforms (**Figure S21**). When applied to extracts of platelet-poor plasma, internal controls showed that high detection specificity required RNA digestion after RCA (**Figure S22a**) to eliminate false-positive signals derived from long RNAs present in extracts (**Figure S22b, S22c**).

**Figure 7.**
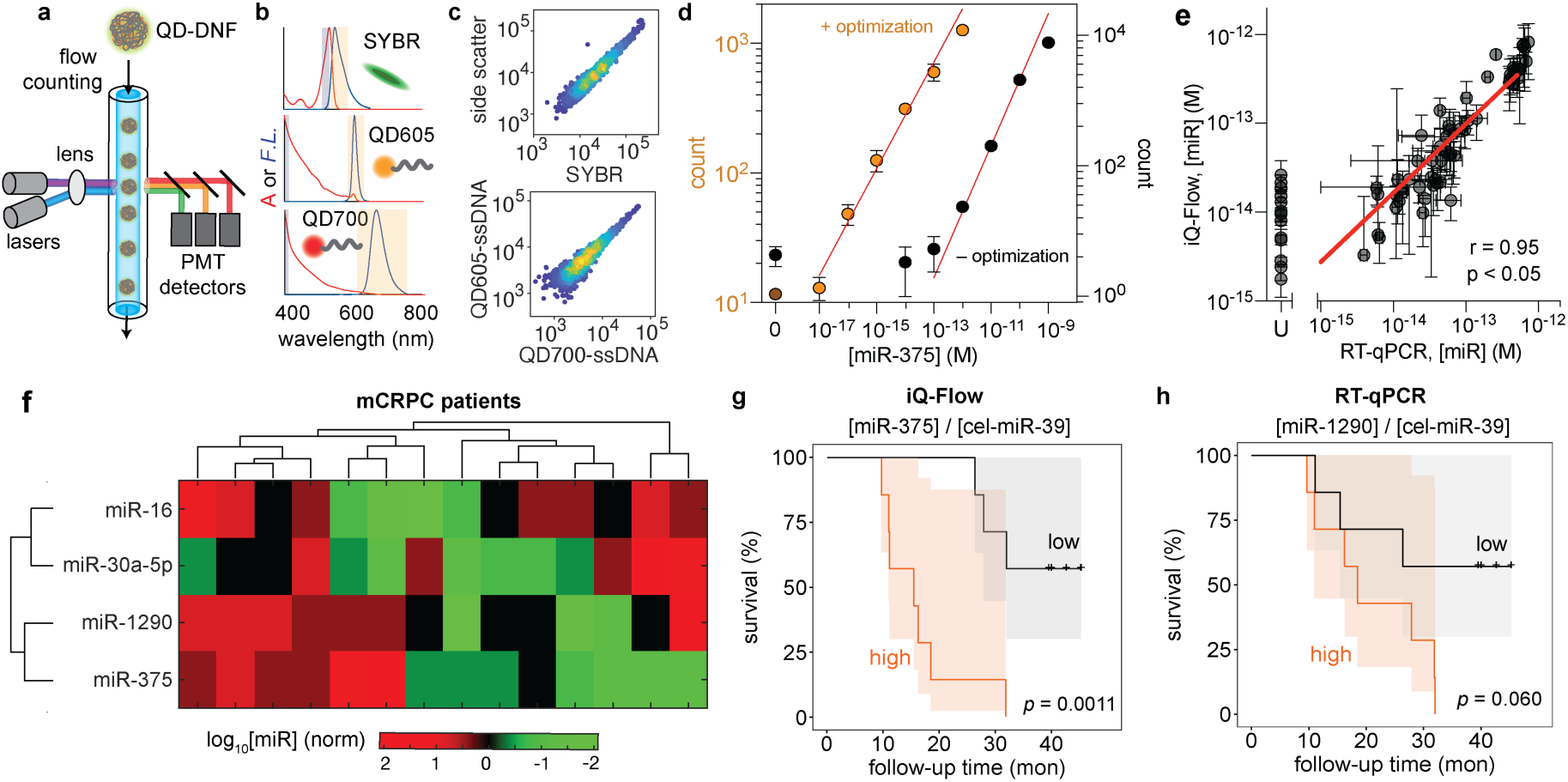
iQ-Flow with flow cytometry readout of miR targets in clinical plasma extracts. **(a)** Schematic shows iQ-Flow counting of fluorescently labeled DNFs from miR targets. **(b)** Fluorophore absorption (A, red) and fluorescence (F.L., blue) spectra for labels based on QDs and dyes with normalized intensities. Highlights indicate emission bandpass filters (orange) and excitation laser band (dark blue). **(c)** Scatter plots show events detected for DNFs labeled with two QD-ssDNA and SYBR. **(d)** Event counts (circles) *versus* miR concentration before (black) and after optimization (orange). Red line indicates the linear regression of the log-log transformed data. Data points and error bars represent mean and S.D., respectively. *N*=3. **(e)** Correlation between iQ-Flow counts and RT-qPCR measurements of 5 biomarkers (cel-miR-39, miR-16, miR-30a-5p, miR-375, and miR-1290) in 14 plasma extracts from mCRPC patients. “U” indicates undetected in the RT-qPCR assay, of which all are miR-375 with biological triplicates. **(f)** Heat map of two prognostic biomarkers (miR-375, miR-1290) and two endogenous normalization controls (miR-16, miR-30a-5p) in 14 mCRPC patient samples. **(g)** Kaplan-Meier curves show a significant survival association with the normalized prognostic biomarkers (miR-375 normalized to cel-miR-39) detected by iQ-Flow. **(h)** Kaplan-Meier curves show survival association with the normalized prognostic biomarkers (miR-1290 normalized to cel-miR-39) detected by RT-qPCR. The orange line and shaded area indicate groups classified as high-risk while the black lines indicate the low-risk group. N = 14.

### iQ-Flow of Clinical Small-Volume Plasma Extracts

In a cohort of 14 mCRPC patients we applied iQ-Flow to quantify prognostic biomarkers miR-1290 and miR-375,^36,52–54^ endogenous normalization controls miR-30a-5p and miR-16,^36,55,56^ and cel-miR-39 spiked in for exogenous normalization.^56–58^ We used small volume samples (50 µL), a challenging clinical biospecimen as extracts cannot be concentrated to overcome assay detection limits. Comparing results with gold-standard RT-qPCR (standard curves in **Figures S23** and **S24**), aggregate agreement was excellent (Pearson’s r = 0.95; *p* < 0.05) with ICC of 0.94 (two-way mixed effects model). Further, iQ-Flow was able to detect miR-375 present in sub- to low-femtomolar levels that were undetectable by RT-qPCR (**Figure S25**). This suggests that iQ-Flow can assist in detecting the biomarkers that are out of or at the edge of the current gold-standard detection techniques from small-volume samples.

### Treatment Failure Time Prediction in mCRPC Patients

Using the iQ-Flow-based miR panel from 14 mCRPC patients normalized by cel-miR-39 (**Figure 7f**), Kaplan-Meier analysis was performed for the two prognostic biomarkers with and without different combinations of normalization controls (**Table S3, S4**). It was found that elevated miR-375 strongly predicted shorter overall survival, to a greater extent than with miR-1290, which is similar to previous findings using large volume blood samples (**Figure 7g** and **Figure S26**).^36^ The predictive power was greater than that using the RT-qPCR-based miR panel, which could only reliably analyze miR-1290 (**Figure 7g-h**), and greater than any other single clinical prognostic factor used for this stage (*e.g.,* prostate-specific antigen [PSA],^59–62^ lactate dehydrogenase [LDH],^63–65^ alkaline phosphatase [ALP],^66,67^ age) (**Figure S27**). A multivariate model for the normalized prognostic miR biomarkers in combination with clinical factors (PSA and ALP) showed a significant association with patient survival (*p* = 0.00039; **Figure S27**). Based on this model, patients with a high clinical factor and high miR-375 had a 2.72-fold higher risk of death than patients with a low-risk score (*p* = 0.004; **Table S5**). This clinical outcome is aligned with previous reports applying RNA sequencing readouts, indicating the reliability of this quantification method for miRs.^36^

## CONCLUSION

Digital assays based on surface pulldown remain limited by inefficient capture and throughput,^7,8,11,16,18^ although creative approaches are being pursued to address this.^12,19,40^ Counting in a flowstream can alternatively eliminate pulldown and increase throughput and readout speed, as is standardized in blood cytology.^1,4,19,20^ Here, in-solution assembly of QD-DNFs from miR targets yields low LOD (47 aM) needed for detecting prognostic miR biomarkers from small-volume plasma or other biofluids, for which sample processing remains a central challenge.^12,25^ The products can be detected well above background signals without wash steps, despite the presence of a high concentration of unbound labels when using spacers^68^ and pH-driven DNF refolding to address the steric hindrance of nanocrystal labels. QDs simplify instrumentation requirements for multiplexed detection of multiple targets^21,69,70^ to allow panels of biomarkers that can improve diagnostic and prognostic value.^71^ Most importantly, low-abundance biomarkers like miR-375 from low-volume plasma extracts could be detected that were outside the range of RT-qPCR, while simultaneously measuring normalization markers with much higher abundance due to a wide dynamic range. The low sample volume required and low LOD may enable the use of blood collected from a finger-prick, while simplifying optical components for point-of-care devices, thereby reducing associated costs. Moreover, the flexibility of designing padlock DNA template and spacer sequences can be applied for the detection of mRNA, ctDNA, or proteins.^1,20,72^ Such a readout platform may lower barriers to the implementation of screening programs in the future.

## METHODS

### RCA template design

A ssDNA padlock probe was designed to contain a complementary sequence to the full-length miRs target, four 20-base probe binding sites for labels with a 5-base spacer for a 180-degree pitch between adjacent QD, and 5’ phosphorylation for ligation (**Figure S1** and **Table S1**). Ligation by SplintR was needed for high specificity RCA-based production of DNFs in comparison with a pre-ligated circular template based on real-time fluorescence intensity measurements (**Figure S28**). The circularization, real-time fluorescence intensity measurement, and characterization of the circularized template are described in the **Supporting Methods**. Multiple template sequences were evaluated based on the previously reported sinusoidal dependence of polymerase activity on template length, and the probe binding site was designed based on the reported improved hybridization specificity with higher C and lower G content.^73–75^ A screening of template lengths between 79 to 174 bases showed a reduction of SplintR ligation efficiency and total DNA synthesis with increasing length, with 121 bases selected as an appropriate balance (**Figure S29**).

### DNF synthesis

The ssDNA padlock probe (100 nM; **Table S1**) was ligated in a 10 µL solution containing synthetic miRs or 10-fold dilution of plasma extracts containing SplintR ligase (0.625 U uL^−1^) in SplintR ligase reaction buffer (pH 7.5, 50 mM Tris-HCl, 10 mM MgCl_2_, 1 mM ATP, 10 mM dithiothreitol, 1.0 U μL^−1^ SUPERase-In RNase Inhibitor, and 1.0 U μL^−1^ Murine RNase Inhibitor at pH 7.5) for 2 h at room temperature. RCA was then initiated through the addition of a 2× polymerase reaction mixture (0.8 U μL^−1^ Φ29 DNA polymerase, 2 mM dNTPs, 0.2 mg mL^−1^ BSA at pH 7.5) and was allowed to proceed for 4 h at 37 °C in a ThermoFisher QuantStudio 3 Real-Time PCR. For quantitative measurement of DNF growth, 0.001% SYBR Gold was added to samples, and its fluorescence signal intervals were monitored for 15 s using LED excitation at 470 nm and emission at 520 ± 10 nm.

Polymerase was then heat-inactivated at 80 °C for 10 min, and the products were stored at −20 °C until further use.

### Conjugation of QDs

QDs coated with an azide-functional polymer (1 µM; QD synthesis, polymer coating, and hydrodynamic size characterization are described in the **Supporting Methods**) were reacted with dibenzocyclooctyne (DBCO)-functionalized ssDNA (**Table S1**) at a 4:1 ssDNA:QD ratio overnight. Excess ssDNA was removed by centrifugal filtration (Amicon Ultra 30 kDa MWCO; six times). Conjugation was verified by electrophoresis, described in **Supporting Methods**, in a 0.5% agarose 2% polyacrylamide mixed gel performed at 120 V for 30 min (**Figure S6**). All procedures were carried out at 4 °C.

### DNF labeling with QDs

DNFs were labeled in a solution containing QD-ssDNA (5 nM unless noted otherwise; **Table S1**) and SYBR Green (0.0316%) for iQ-Flow, spacers (0.5 nM unless noted otherwise; **Table S1**), BSA (0.5% for in-solution imaging, 0.03% for flow cytometry), and MgCl_2_ (1.5 mM) in Britton–Robinson buffer (BrB buffer; 40 mM H_3_BO_3_, 40 mM CH_3_COOH, 40 mM H_3_PO_4_, and 100 mM NaCl, pH 7.4). DNF samples were first mixed with spacers and BSA in BrB buffer, and denatured at 80 °C for 10 min. For reactions including pH-driven reassemby, the pH of the solution was then raised (pH = 10.4 unless otherwise noted) with 0.1 M NaOH, and QD-ssDNA was added at room temperature. After 10 min, the pH was reduced to 7.4 with 0.1 M HCl, and the mixture was incubated for 4 h at 37 °C. MgCl_2_ was then added to stabilize the QD-DNF complex. For multiplexed measurements, the concentrations of QD-ssDNA and spacers were adjusted to match fluorescence intensities with excitation at 405 nm wavelength: 50 nM for QD445–R1’, L1– R1, QD525–R2’, L2–R2, QD570–R3’, and L3–R3, 10 nM for QD605–R4’ and L4–R4, and 2.5 nM for QD625–R5’ and L5–R5. After labeling, DNFs were diluted 3.3-fold in glycerol for in-solution imaging or used undiluted for flow cytometry.

### DNF labeling with dyes

DNFs were labeled using the same method as with QDs, except dye-ssDNA (**Table S1**) was used in place of QD-DNA, and no spacers and pH-driven reassembly were included. For on-surface assays, the mixture contained biotin–L3 (100 nM) and Alexa546–L1 (100 nM) with 2% BSA in phosphate-buffered saline (PBS). For in-solution assays, the mixture contained Alexa546–L1 (100 nM) with 0.5% BSA in PBS. The mixture was incubated for 4 h at room temperature, followed by imaging.

### In-solution assays

For data in **Figure 2a, 2c, 2e,** and **Figure 3**, DNFs labeled with Alexa546–L1 or QD605–L1 were diluted in glycerol (70%) to prevent motion-blur during imaging. The blocking solution was empirically tuned in **Figure S30**. Fluorescence micrographs were acquired on a 50-well chambered coverglass at an image plane above the surface using a Zeiss 710 confocal scanner with an Azio Observer Z1 inverted microscope stand with 63× objective (NA 1.4) and 405-nm or 561-nm wavelength laser excitation. For non-spectral analysis, emission intensity was measured at 530 nm and 727 nm wavelengths. For multispectral imaging, a 26-channel spectral range between 442 nm and 650 nm with a 9.8 nm interval was acquired using a QUASAR 34-channel spectral detector.

### On-surface assay

For data in **Figure 2b, 2d, and 2f**, surface-based assays were performed using previously reported protocols using streptavidin-functionalized PEGylated substrates.^8,16^ 50-well chambered coverglass was cleaned with potassium hydroxide (1 M) with sonication for 10 min. Surfaces were then cleaned with oxygen plasma and immediately incubated in a solution of methanol (93.46%), glacial acetic acid (4.67%), and 3-aminopropyltriethoxysilane (1.87%) for 10 min at room temperature in the dark, followed by 1 min sonication, and an additional 10 min incubation. Afterward, a freshly prepared solution containing monomethoxy monosuccinimidyl ester poly(ethylene glycol) (mPEG5000-NHS, 2.375% w/v) and monobiotin monosuccinimidyl ester poly(ethylene glycol) (biotin-PEG5000-NHS, 0.125% w/v) in sodium bicarbonate (10 mM) was applied to the surfaces, which were incubated in a humidified chamber for 4 h. Between each step, the coverglass was washed with water twice and dried with a nitrogen flush. After functionalization, streptavidin (0.2 mg/mL) in PBS was added for 10 min, followed by two PBS washes. Dye-DNFs labeled with biotin–L3 and Alexa546–L1 (diluted 3.33-fold to match concentrations used in the in-solution assay) were incubated on the streptavidin-functionalized coverglass for 4 h at 25 °C. The substrates were then washed twice with PBS and imaging immediately with HiLo illumination on a Zeiss Axio Observer.Z1 inverted microscope with a 100× objective (NA 1.45) and 561 nm wavelength diode laser excitation with 564 nm/15 nm bandpass filter (Semrock). Images were collected with a Photometrics EXcelon Evolve 512 EMCCD camera after light was passed through a 600 nm/37 nm bandpass filter (Semrock).

### Quantification of labels per DNF

To quantify the number of dyes or QDs per DNF, DNF intensities from dynamic micrographs of labeled DNFs in solution were compared with intensities of single dyes and QDs adsorbed to a surface. DNFs labeled with Alexa546-L1 or L1 conjugates of different QDs (**Table S1**) were imaged in-solution with HiLo illumination on a Zeiss Axio Observer.Z1 inverted microscope with a 100× objective (NA 1.45). Laser excitation sources were 561 nm for dyes and 405 nm for QDs, and 200 frames were acquired at 4.35 frames per second using a Photometrics EXcelon Evolve 512 EMCCD camera using Zeiss ZEN software with parameters listed in **Figure S8**. Excitation light was filtered through bandpass filters (390 nm/40 nm or 564/15 nm; Semrock). Emission signals were filtered using bandpass filters (525 nm/50 nm, 562 nm/40 nm, 600 nm/37 nm, or 732 nm/68 nm; Semrock), dependent on the emission wavelength of the label. For imaging of individual immobilized labels, dyes and QDs diluted to 0.01 nM in PBS were drop-cast on glass coverslips and immediately rinsed with PBS to remove unbound labels before imaging using identical parameters as labeled DNFs in solution (**Figure S8**).

### Image analysis

Micrographs as 8-bit uncompressed TIFF files were used for spot counting using the MATLAB Multi-target Tracking (MTT) algorithm to determine the location and intensity of each spot.^38^ In the algorithm, spot detection is performed by evaluating 9×9 windows in the image using a generalized likelihood ratio test to decide if a spot fits a point spread function, assuming Gaussian noise. The algorithm then subtracts each spot and repeats the analysis until all spots are detected. The threshold applied for the hypothesis test is normalized as the probability of false positives per 512×512 pixel image (false positives per ∼250,000 windows). Time-course image stacks were processed using previously reported MATLAB scripts to obtain the relative brightness *B*_rel_ of each trajectory, calculated as the mode of the maximum 6% of the brightness distribution, using trajectories with a minimum length of 10 frames.^49^ For multispectral analysis, spectral image stacks were analyzed as if they were time-course image stacks, and spots were connected across a spectral sliding window of 26 channels to acquire the full fluorescence spectrum of identified spots from 442 nm to 650 nm. The spectrum from each spot was then used to fit with a Gaussian function to determine the peak and full width at half-maximum of the emission wavelength, with aggregated data across spots summarized in **Table S2**.

### QD-DNF characterization

TEM micrographs were collected with a JEOL 2100 CRYO electron microscope, and SEM micrographs were collected with a Hitachi S4800 electron microscope, both in the Frederick Seitz Materials Research Laboratory Central Research Facilities at the University of Illinois. For TEM, each sample was prepared by drop casting a sample suspension on an ultrathin carbon-coated Cu-TEM grid before drying the solution in ambient atmospheric conditions. For SEM, samples were purified and drop-cast onto a cleaned silicon wafer. The samples were then dried at 60 °C overnight and sputter-coated with gold for 40 s before imaging.^47^ The surface area of each object was quantified using ImageJ. Images were first converted to 8-bit grayscale, and an appropriate threshold was applied to segment the objects from the background. Gaussian fitting of the log-transformed data was then used to calculate the mean surface area. Fluorescence spectra were acquired using a NanoLog Horiba Jobin Yvon with Fluo Essence V3.5 software (HORIBA Scientific). Absorption spectra were collected using a Cary series UV-Vis-NIR spectrophotometer with Cary Win UV Scan Application Version 6.00 1551 software (Agilent Technologies). FCS data were acquired and analyzed as previously reported.^48,49^ Briefly, samples were diluted to 10 nM in PBS and transferred to 8-well glass-bottom LabTek chambers. Fluorescence time scans were collected using an ISS Alba FCS instrument with 470 nm diode laser excitation and a single-photon avalanche photodiode detector. Traces were collected for 10 s at 10^5^ Hz.

### iQ-Flow with flowstream readout

Labeled DNFs were transferred to 5 mL round-bottom polystyrene tubes for loading into a BD LSR Fortessa Flow Cytometry Analyzer or BD Symphony Flow Cytometry Analyzer. All acquisition parameters were tested prior to each experiment using Flow-Check Fluorospheres to confirm that emission parameters were within expected intensity ranges. All events were recorded for which intensities were greater than the minimum detection threshold of 200 r.f.u. for side scattering and SYBR Green. Data for 11.67 μL were collected for 20 s. Fluidics were cleaned with desalinated water to remove residual samples between experiments. Data were analyzed and visualized with FlowJo software (version 10.8.1; BD Bioscience System & Reagent, Inc.). The gating strategy followed that of our previous manuscript.^4^

### Patient specimens

mCRPC patients who received standard cancer treatment were recruited with consent for research biospecimen collection under an institutional IRB-approved study of “Total Cancer Care” (*IRB# 00089989*; *IRB# 00139755*). Uniform blood collection was then performed under Clinical Laboratory Improvement Amendments supervision at Huntsman Cancer Institute for uniform processing. Briefly, the whole blood was collected in 4.5 mL Vacutainer tubes. On the day of collection, platelet-poor plasma was separated by centrifugation at 2000 rpm for 10 min at 4 °C, and a second centrifugation of the supernatant at 3000 rpm for 10 min. The supernatant was transferred to a sterile tube, leaving approximately 1 cm above the buffy coat to avoid contamination with white blood cells and platelets. Plasma was aliquoted into 500 µL volumes, visually inspected for hemolysis, and frozen at −80 °C until further use. Granular clinical annotation was obtained from electronic medical records (EMRs) for survival outcomes.

### RNA extraction

Exosomes from plasma were isolated using ExoQuick reagents. Briefly, ExoQuick (60 µL) was added to plasma (50 µL), and exosomes were precipitated overnight at 4 °C. Samples were then centrifuged at 1500 *g* at 4 °C for 30 min. Aqueous phase total RNA was extracted from isolated exosomes using TriZol, and 44 attomoles of cel-miR-39 spike-in was added to the solution to establish extraction efficiency. The collected aqueous phase was diluted into a solution containing ethanol (64.5% v/v), 96.8 mM sodium acetate, and ∼8 µg/mL glycogen for RNA precipitation overnight at –20 °C. RNA was then sedimented through centrifugation (10,000 *g* for 10 min at 4 °C), and the pellet was resuspended in 75% ethanol and precipitated again by centrifugation (7500 *g* for 5 min at 4 °C). The pellet was dissolved in RNase-free water (40 μL) and stored at −80 °C until analysis.

### RT-qPCR

Commercial TaqMan miR assays were used to quantify the targets, controls, and spike-in miR from total RNA extracts. All assays were independently optimized to achieve a dynamic range of detection between 1 pM and 1 fM, ensuring a broad dynamic range and high sensitivity to quantify low-concentration miRs accurately. Standard curves were generated for each miR using synthetic miR standards for miR-375, miR-1290, miR-30a-5p, miR-16, and cel-miR-39 (**Table S1**) and used to determine concentrations of test samples from Ct values. Total RNA extract from plasma was diluted 10-fold before the RT reaction to minimize enzyme inhibition in the PCR assays.

### Survival analysis

The survival time was recorded from the time of plasma collection after androgen deprivation therapy failure to death or last follow-up. Association between the detected miR level and overall survival was assessed using Kaplan-Meier survival curves, with statistical significance determined by the log-rank test. Hazard ratios and 95% confidence intervals were estimated using univariable and multivariable Cox proportional hazards regression models. For the multivariate analysis, the risk score was calculated by a linear combination of normalized and standardized miR level and clinical factors like PSA and ALP, weighted by their estimated regression coefficients. All analyses were performed in R (version 4.2.1) using the *survival* package and visualized with the *survminer* and *ggplot2* packages.

### Statistical analysis

All statistical analyses, unless stated otherwise, were performed in Prism 9.0 software (GraphPad). The intraclass agreement test was performed in R (version 4.2.1) using the *psych* package.

## Data Availability

All data produced in the present work are contained in the manuscript

## Acknowledgements

This work was supported by funds from the National Institutes of Health (R01CA227699, R01EB032725) and the Cancer Center at Illinois (CCIL).

## Author contributions

C.-W.K. and A.M.S. conceptualized the assays, designed experiments, and developed the protocols. S.S. and W.L. performed QD synthesis. C.-W.K. performed QD processing and characterizations, all iQ-Flow procedures, machine learning guided assay optimization, and FCS measurements. M.K. recruited the patient cohort and collected whole blood. M.K., A.M.S., and L.W. designed the biomarker selection. S.N. performed RNA extraction from patient plasma and RT-qPCR. C.-W.K. performed image analysis, statistical analysis, and survival analysis. C.-W.K. and A.M.S. wrote the initial manuscript draft. All authors contributed to manuscript edits.

