## Supporting Information for "Quantum Dot Encoding for In-Solution Single-Molecule Biomarker Counting in Metastatic Prostate Cancer"

### Table of Contents

1. Supporting Figures ……………………………………...S2
2. Supporting Tables………………………………………S32
3. Supporting Materials and Methods ……..……………S35
4. References……….……..…………………………...S40

### 1. Supporting Figures

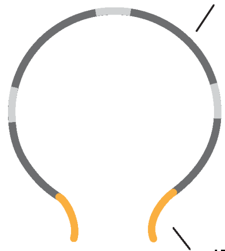


**Figure S1**. **Schematic of ssDNA padlock probe design.** The miR target region (yellow) comprises two ~11-base sequences that endow miR specificity. Four 20-base labeling regions (dark grey) allow labeling with label-ssDNA or spacers (**Table S1**), separated by a 5-base junction (light grey) for 180˚ pitch between adjacent labels.

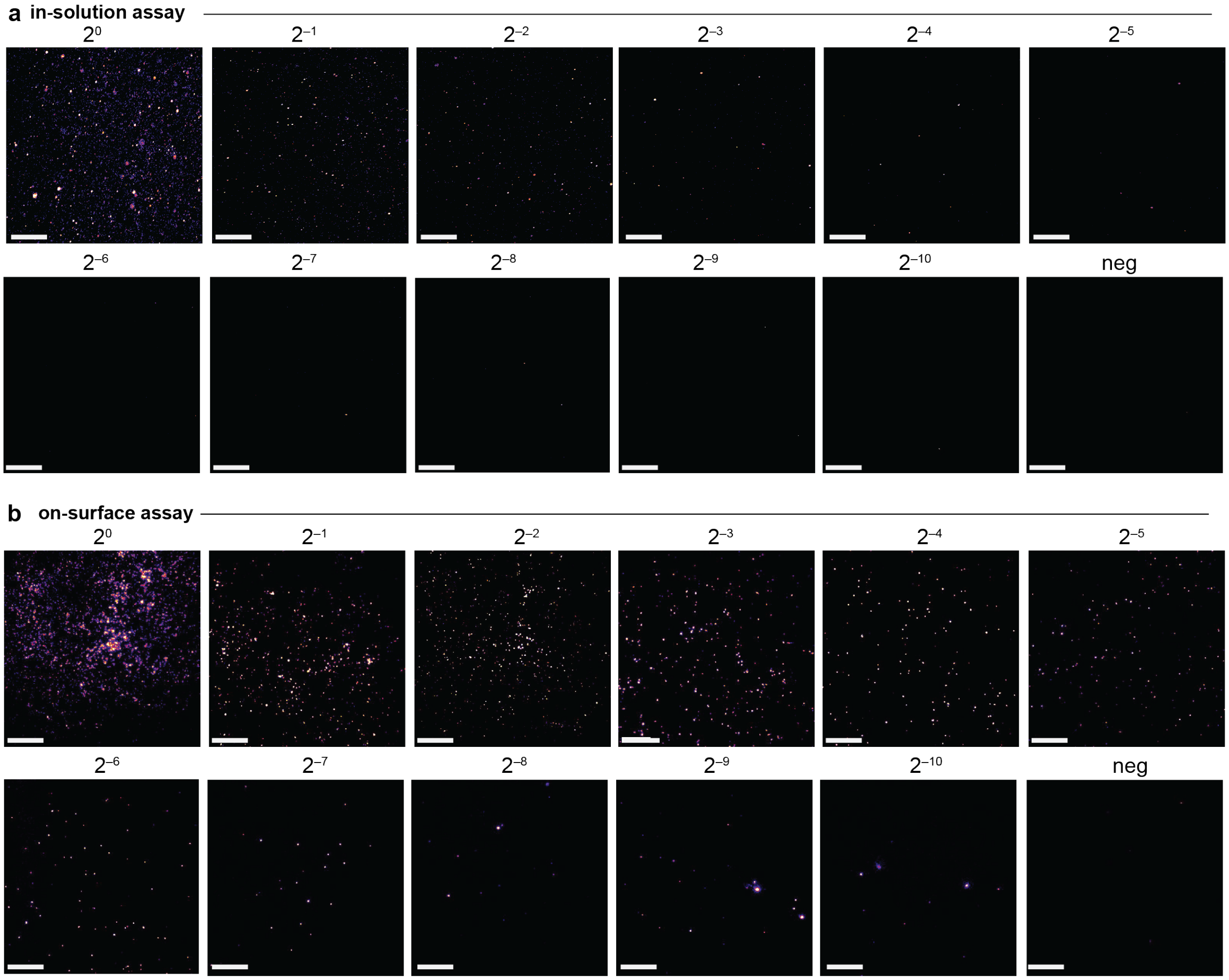


**Figure S2**. **Representative uncropped fluorescence micrographs of DNFs** at 2-fold serial dilutions for **(a)** the in-solution assay and **(b)** the on-surface assay. The number above the micrograph indicates fold dilution from a 1.5 nM solution of Alexa546-DNFs. “neg” indicates samples contain no DNF. Scale bar: 20 μm.

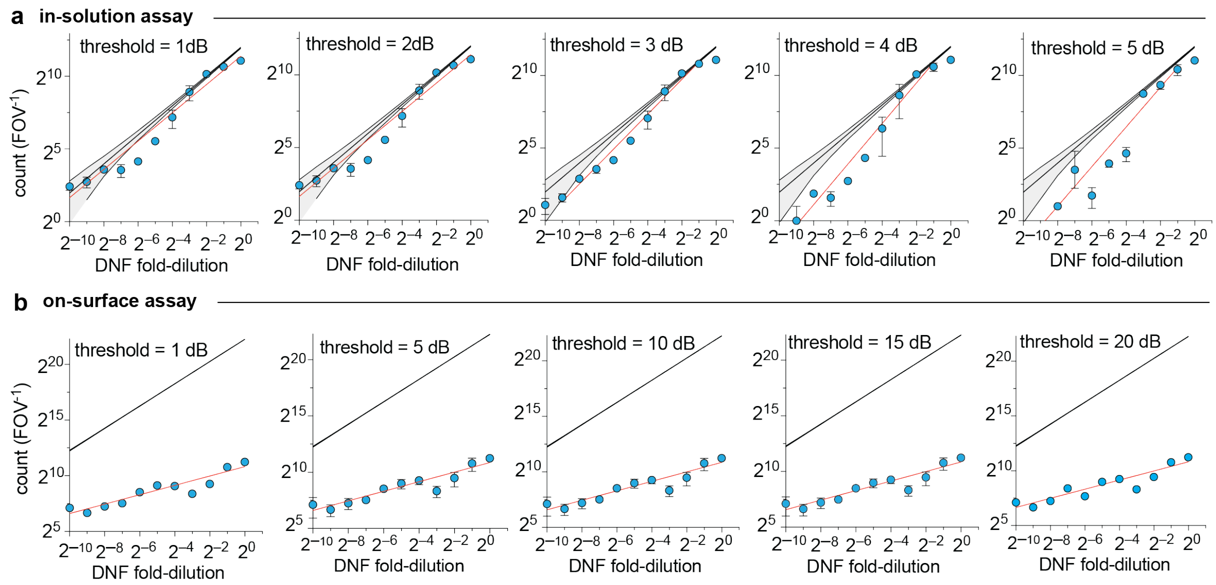


**c in-solution assay**

| threshold (dB) | slope | $R^2$ |
|---|---|---|
| 1 | 0.98 | 0.64 |
| 2 | 0.98 | 0.64 |
| 3 | 1.2 | 0.97 |
| 4 | 1.4 | 0.95 |
| 5 | 1.4 | 0.92 |

**d on-surface assay**

| threshold (dB) | slope | $R^2$ |
|---|---|---|
| 1 | 0.45 | 0.81 |
| 5 | 0.45 | 0.81 |
| 10 | 0.45 | 0.76 |
| 15 | 0.45 | 0.82 |
| 20 | 0.42 | 0.76 |

**Figure S3**. **Dependence of in-solution and on-surface assay metrics on spot detection threshold.** Data correspond to **Figure 2** and **S2**. **(a)** Dependence of spot count per field of view (FOV) on DNF concentration for the in-solution assay with indicated thresholds from 1 to 5 dB, with error bar indicating standard deviation of nine technical replicates. The black solid line indicates the theoretical count number derived from the absolute DNF number in the confocal volume. The red line represents a linear regression of the log-log-transformed data. **(b)** Dependence of spot count per FOV on DNF concentration for the on-surface assay with indicated thresholds from 1 to 20 dB, with error bars indicating standard deviation of nine technical replicates. The black solid line indicates the theoretical count number derived from the absolute DNF number in the total sample volume, with a grey shading area background indicating the confidence interval. The red line represents a linear regression of the log-log-transformed data. **(c)** Slope and goodness of fit ($R^2$) for data in panel (a). **(d)** Slope and $R^2$ for data in panel (b). Both assays showed a saturated signal for samples with dilution factors between $2^0$ and $2^1$ because the concentrated DNF leads to a dense field of spots, for which single spots cannot be fully discriminated (**Figure S1**). Additionally, absolute values in the in-solution assay were more dependent on the spot detection threshold applied in the detection algorithm (Pearson correlation test, r = 0.93) due to the presence of unbound labels.

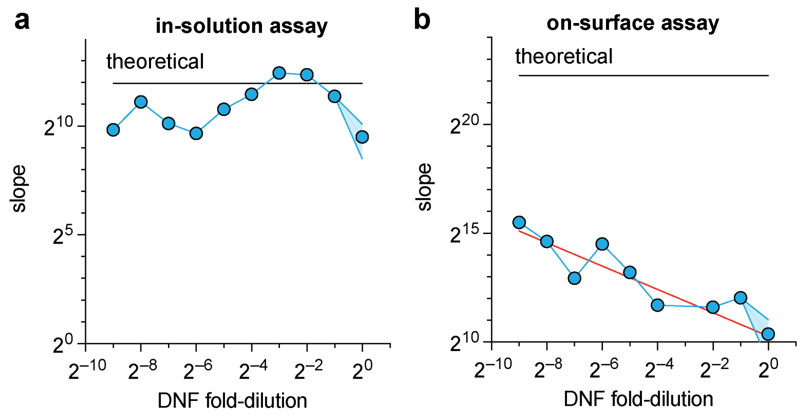


**Figure S4. Count slope *versus* DNF dilution factor for data in Figure 2**. Slope was calculated as the count difference between consecutive data points divided by the difference in dilution factors. Plots show **(a)** in-solution assay and **(b)** on-surface assay. Points indicate the mean, and the shaded area indicates the standard deviation for 9 technical replicates. The black line indicates the linear slope of the theoretical value of an absolute assay. The red line in (b) represents a linear regression of the on-surface assay across all dilutions after log-log transformation of data (slope = –0.48; $p$ = 0.0008).

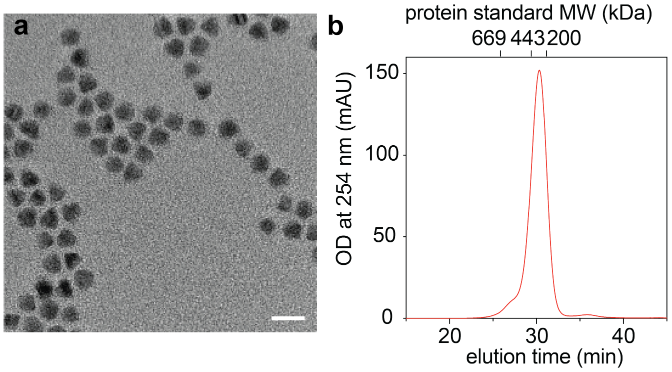


**Figure S5. QD605 structure**. **(a)** Transmission electron micrograph of core/shell CdSe/CdZnS nanocrystals used in this work. Scale bar: 10 nm. **(b)** Gel permeation chromatogram (GPC) of the nanocrystals after coating with azide-functional multidentate polymer, showing an elution time corresponding to globular protein size standards between 200 and 443 kDa molecular weight (MW). OD = optical density. mAU = milli-absorbance unit.

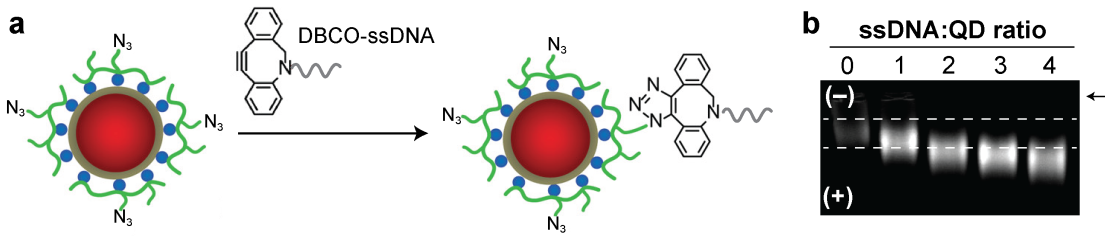


**Figure S6. ssDNA conjugation to QDs. (a)** Schematic showing azide-functionalized QD reaction with ssDNA functionalized with dibenzylcyclooctyne (DBCO) to generate a covalent triazole-linked conjugate. **(b)** Agarose-polyacrylamide gel electrophoresis of QD-ssDNA conjugates with reactions performed at the indicated molar ratios. The white horizontal dashed line indicates the location of the QD-only position in the gel. The arrow indicates the position of the sample loading well in the gel. Positive and negative signs indicate directions of electrode polarity. For a 4:1 ssDNA:QD ratio, nearly all QDs were functionalized with at least 1 ssDNA indicated by complete migration outside the dashed lines.

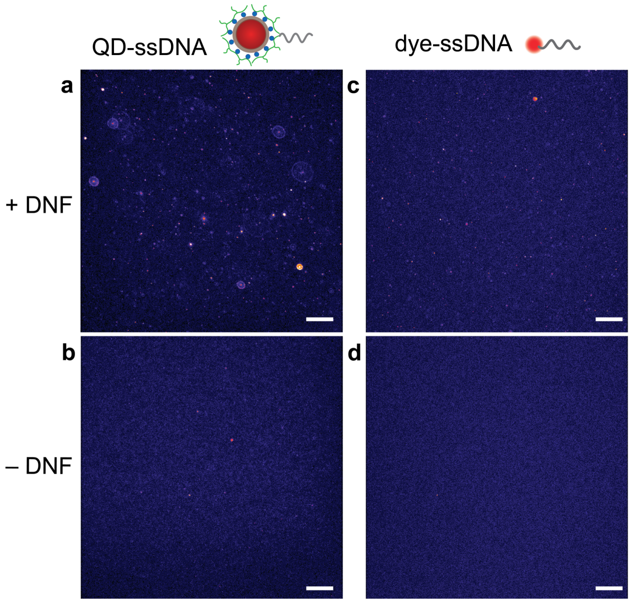


**Figure S7. Uncropped fluorescence micrographs of DNFs labeled with QD-ssDNA or dye-ssDNA from Figure 3.** Panels show **(a,c)** labels with DNFs or **(b,d)** labels alone. Scale bar: 20 μm.

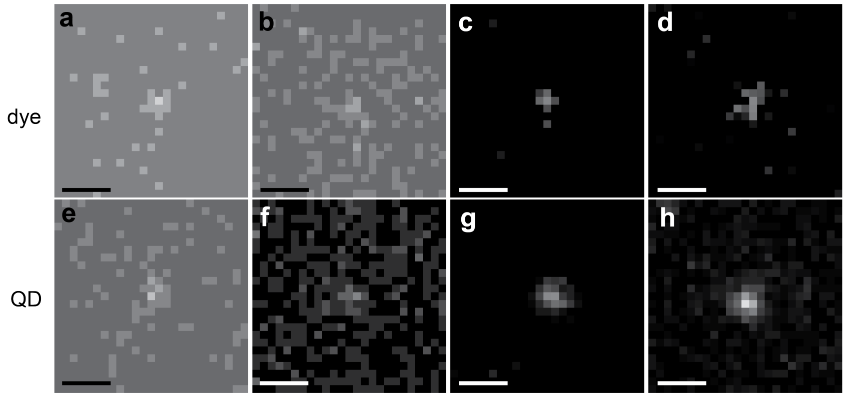


**Figure S8. Single dye and QD immobilized on glass coverslips, imaged at different concentrations and different conditions.** Data were used for calibration of DNFs labeled with each fluorophore type. A single dye or QD is shown near the center of each image, excited with a 100 mW 561-nm laser (dye) or a 100 mW 405-nm laser (QD). For each, the laser power (*P*), camera exposure time (*t*), and electron-multiplying (EM) gain were tuned. **(a)** *P* = 25%, *t* = 25 ms, EM = 100. **(b)** *P* = 25%, *t* = 25 ms, EM = 400. **(c)** *P* = 40%, *t* = 200 ms, EM = 400. **(d)** *P* = 70%, *t* = 200 ms, EM = 500. **(e)** *P* = 20%, *t* = 25 ms, EM = 100. **(f)** *P* = 25%, *t* = 25 ms, EM = 400. **(g)** *P* = 40%, *t* = 200 ms, EM = 500. **(h)** *P* = 70%, *t* = 200 ms, EM = 500. Scale bar: 1 μm.

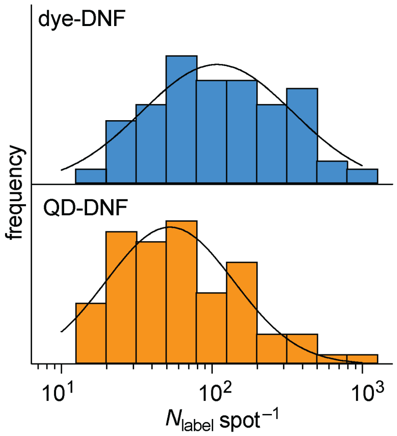


**Figure S9. Number of labels per DNF corresponding to data in Figure 4b of 100 nM labels**. Dye-DNFs are shown in blue, and QD-DNFs are shown in orange. The solid black lines show a fitted single Gaussian function after log-log transformation of data ($R^2$ = 0.86 for dye-DNF, and $R^2$ = 0.83 for QD-DNF). $N$ = 3.

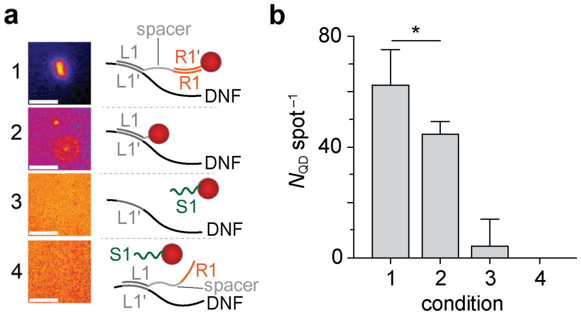


**Figure S10**. **Engineered nucleic acid spacers reduce the steric hindrance of QD for DNF labeling. (a)** Impact of nucleic acid spacers on the number of QD (*HD* = 13.4 nm) per spot showing representative micrographs of labeled DNFs. Experimental conditions shown are numbered as follows. *1*: Spacer L1–R1 is complementary to DNF (L1') and QD-ssDNA (R1'); *2*: QD-ssDNA (L1) is complementary to DNF (L1') without spacer; *3*: QD-ssDNA (scramble sequence S1)is not complementary to DNF; *4*: Spacer L1–R1 is complementary to DNF (L1') but not QD-ssDNA (S1); Scale bar: 5 µm. Sequences are in **Table S1**. **(b)** QDs per DNF spot with experiment conditions described in (a). (mean ± S.D., $N$ = 3). * indicates $p < 0.05$.

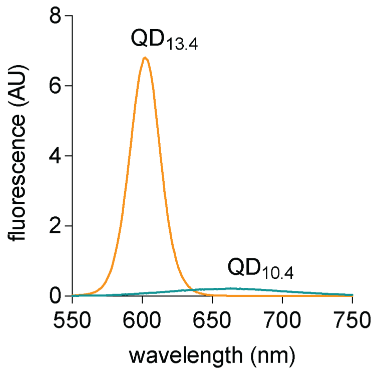


**Figure S11. Fluorescence spectra of $QD_{13.4}$ (orange line) and $QD_{10.4}$ (green line).** Both QDs had the same concentration and were excited with 405-nm wavelength light at the same power. AU = arbitrary units.

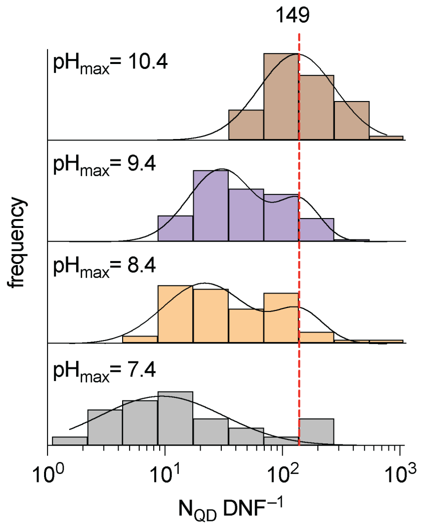


**Figure S12. QDs per DNF measured for QD-DNFs after labeling through pH-switching to $pH_{max}$ followed by return to pH 7.4.** The solid black lines show fitted single or two-component Gaussian functions ($R^2$ = 0.67 for $pH_{max}$ = 7.4, 0.97 for $pH_{max}$ = 10.4). The red dashed line indicates the mean number of QD per DNF for $pH_{max}$ = 10.4 (149 QD $spot^{-1}$).

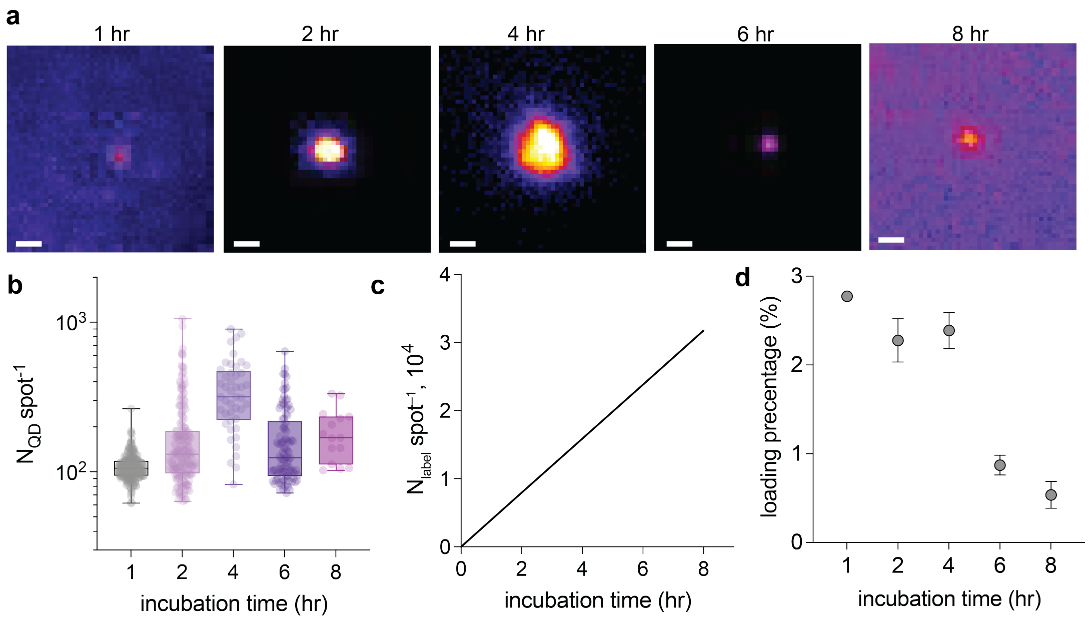


**Figure S13. Dependence of spot counting fidelity on RCA reaction time. (a)** Representative fluorescence micrographs of DNFs with different RCA reaction times after QD labeling and pH-driven reassembly. Scale bar = 0.5 µm. **(b)** Number of QDs per spot after the indicated RCA reaction time. Boxes indicate 25/75 percentiles with the enclosed line indicating the mean. Whiskers are the maximum and minimum values. **(c)** Estimated number of label regions per DNF assuming Φ29 DNA polymerase synthesizes 2000 nt per min.[1] **(d)** The estimated QD loading percentage is the ratio of $N_{QD}$ to $N_{label,}$ showing a marked reduction when the reaction time exceeds 4 h, which may be due to depletion of QD-ssDNA label from the solution. Increasing QD-ssDNA concentration may further enhance labelling, but would be impractical for the assay. The error bar represents S.D. for $N$ = 3.

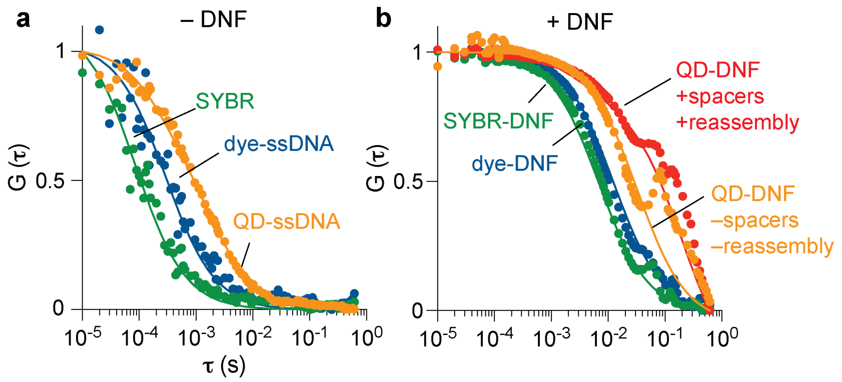


**c**

| label class | DNF | spacers and pH reassembly | *HD* (nm) |
|---|---|---|---|
| SYBR | – | – | 0.480 ± 0.070 |
| | + | – | 106 ± 28 |
| dye-ssDNA | – | – | 3.82 ± 1.02 |
| | + | – | 146 ± 38 |
| QD-ssDNA | – | – | 13.9 ± 1.0 |
| | + | – | 520 ± 68 |
| | + | + | 1330 ± 110 |

**Figure S14. Hydrodynamic sizes of DNFs and labels by FCS.** Representative normalized autocorrelation functions are shown for **(a)** labels at 10 nM, showing dye-ssDNA (blue), SYBR Gold intercalating dye (green), and QD-ssDNA (orange), and **(b)** DNFs at 50 pM, showing those labeled with dye-ssDNA (blue), SYBR (green), QD-ssDNA without spacers and pH-driven reassembly (orange), and QD-ssDNA with spacers and pH-driven reassembly (red). Solid lines indicate fitting curves of the autocorrelation curve. Points represent the mean of five technical replicates. **(c)** Summary of calculated hydrodynamic diameters (*HD*), with standard deviation calculated from five technical replicates.

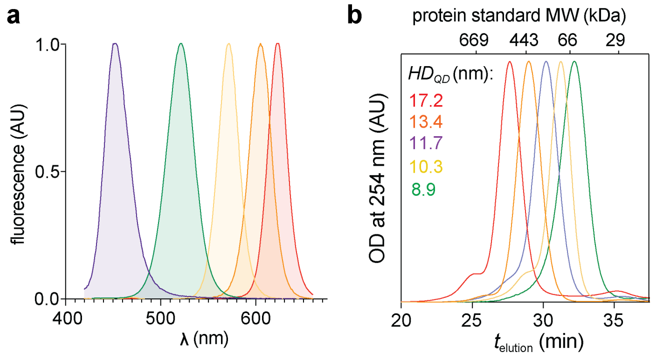


**Figure S15. Spectra and sizes of a color series of core/shell QDs**. Nanocrystals are composed of core/shell CdZnSe/CdZnS for QD445 (purple) or CdSe/CdZnS for QD525 (green), QD570 (yellow), QD605 (orange), and QD625 (red). **(a)** Fluorescence spectra. **(b)** Gel permeation chromatograms with indicated median hydrodynamic diameters shown as calculated using protein size standards shown on the top x-axis. AU = arbitrary units. OD = optical density.

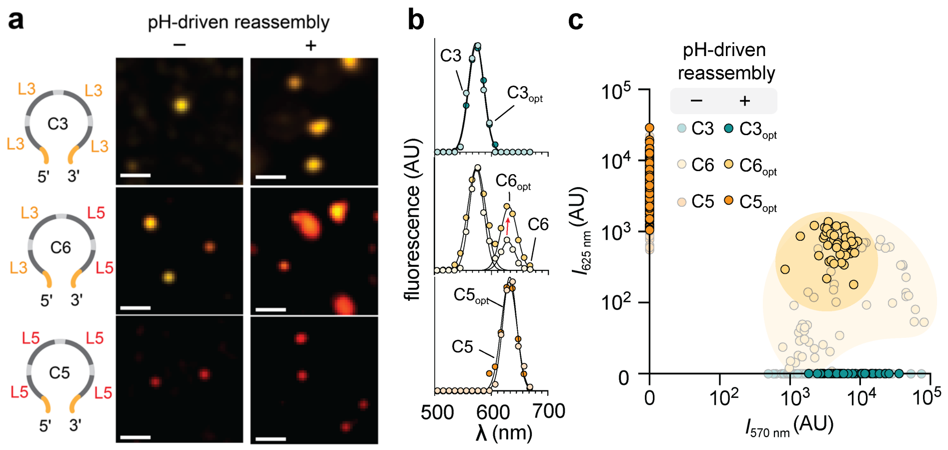


**Figure S16. Two-channel fluorescence intensities for ratiometrically labeled QD-DNFs corresponding to data in Figure 6g-i. (a)** Representative multispectral micrographs of ratiometrically barcoded QD-DNFs with a schematic showing in-solution multiplexed counting of QD-DNFs using ratiometric barcoding with templates containing labeling regions L3 and L5 in different numbers (4:0, 2:2, and 0:4). Scale bars = 5 μm. **(b)** Representative fluorescence spectra of detected spots from panel (a), comparing without pH-driven reassembly and with pH-driven reassembly (indicated with opt). A Gaussian fit (black) is shown for each intensity-normalized spectrum. **(c)** Two-color fluorescence intensities (*I*) of DNFs deriving from barcodes with single-label sequences (C3, C5) or two-label sequences (C6). The yellow shaded area shows the range of fluorescence intensities of dual-labeled DNFs with spacers and pH-driven reassembly (indicated with opt) or without spacers and pH-driven reassembly. The distribution of relative intensities becomes more uniform with the optimized procedure. AU = arbitrary units.

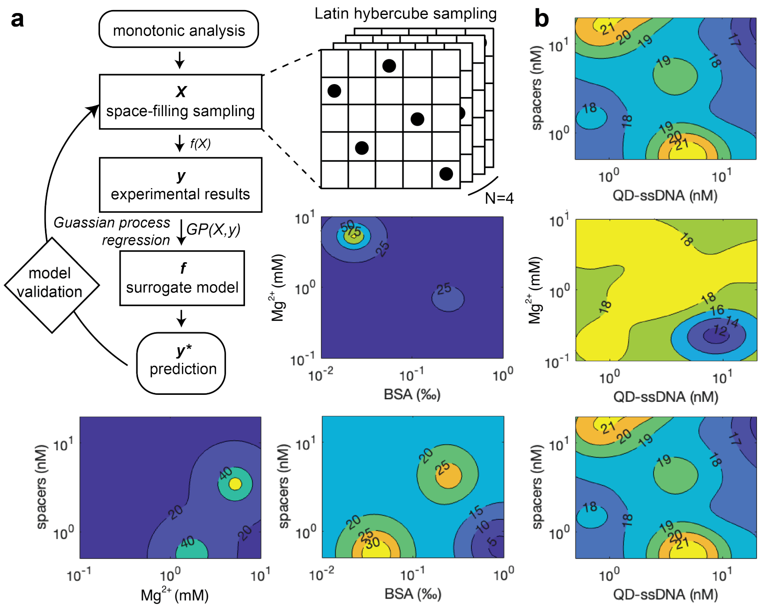


**Figure S17. Machine learning-based iQ-Flow assay optimization. (a)** Surrogate modeling workflow. First, single-parameter (monotonic) analysis was performed based on the concentration of QD-ssDNA, spacers, magnesium (II) ion ($Mg^{2+}$), and 3 blocking agents (yeast tRNA, dextran sulfate, and bovine serum albumin; BSA), as shown in **Figure S18**. A Latin hypercube experimental design with an evolutionary operation was then applied to identify the most space-filling training group (*N* = 25) across four selected parameters, including QD-ssDNA, spacers, $Mg^{2+}$, and BSA, by iteratively mutating previous results to achieve the optimal solution. This was followed by Gaussian Process Regression (GPR) to predict the surrogate model based on the experimental results to solve the confounding effects between the parameters. An extra testing group (*N* = 9) was then applied to evaluate the prediction accuracy of the regression model. **(b)** The surrogate model informed the trajectory of the assay signal-to-background ratio (SBR) of counts, showing predicted hotspots in the SBR of event count trajectories. The SBR trajectories shows a distinct hotspot region for optimal assay performance, where the experimental parameter combinations: 5 nM QD-ssDNA, 0.5 nM spacers, 0.05% BSA, and 1.5 mM $Mg^{2+}$.

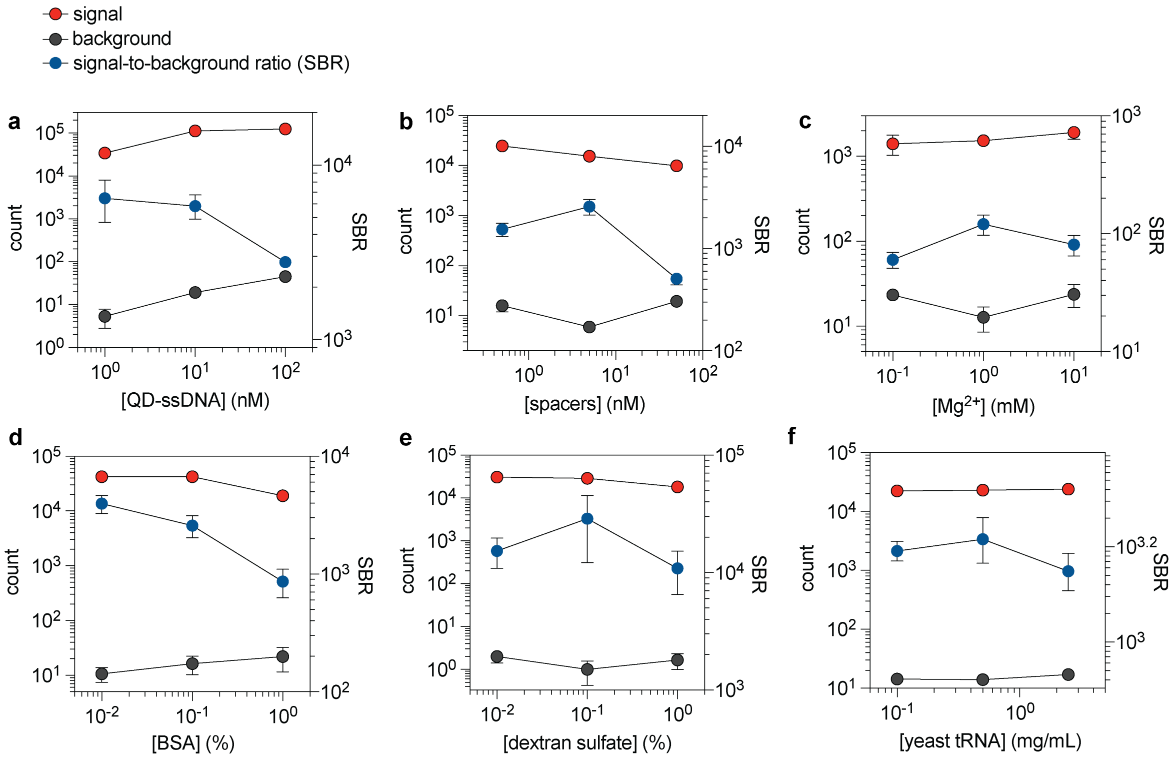


**Figure S18. Estimations for Latin hypercube design using monotonic trends for parameters expected to change the assay LOD**. Plots show signal (50 pM DNF, red circle), background (0 pM DNF, gray circle), and the SBR (blue circle) with respect to the independently tuned parameter. Parameters include concentrations of **(a)** QD-ssDNA, **(b)** spacers, **(c)** $Mg^{2+}$, **(d)** BSA, **(e)** dextran sulfate, and **(f)** yeast tRNA. Error bars represent the S.D. from three technical replicates. Both QD-ssDNA and spacers are expected to modulate the number of QDs per DNF and the amount of unbound QD-ssDNA in the ambient solution. BSA reduces the nonspecific binding of QDs but also can induce the formation of QD aggregates at higher concentrations. $Mg^{2+}$ stabilizes the DNA duplex but destabilizes QDs at higher concentrations. Dextran sulfate and yeast tRNA both prevent nonspecific binding of nucleic acids.

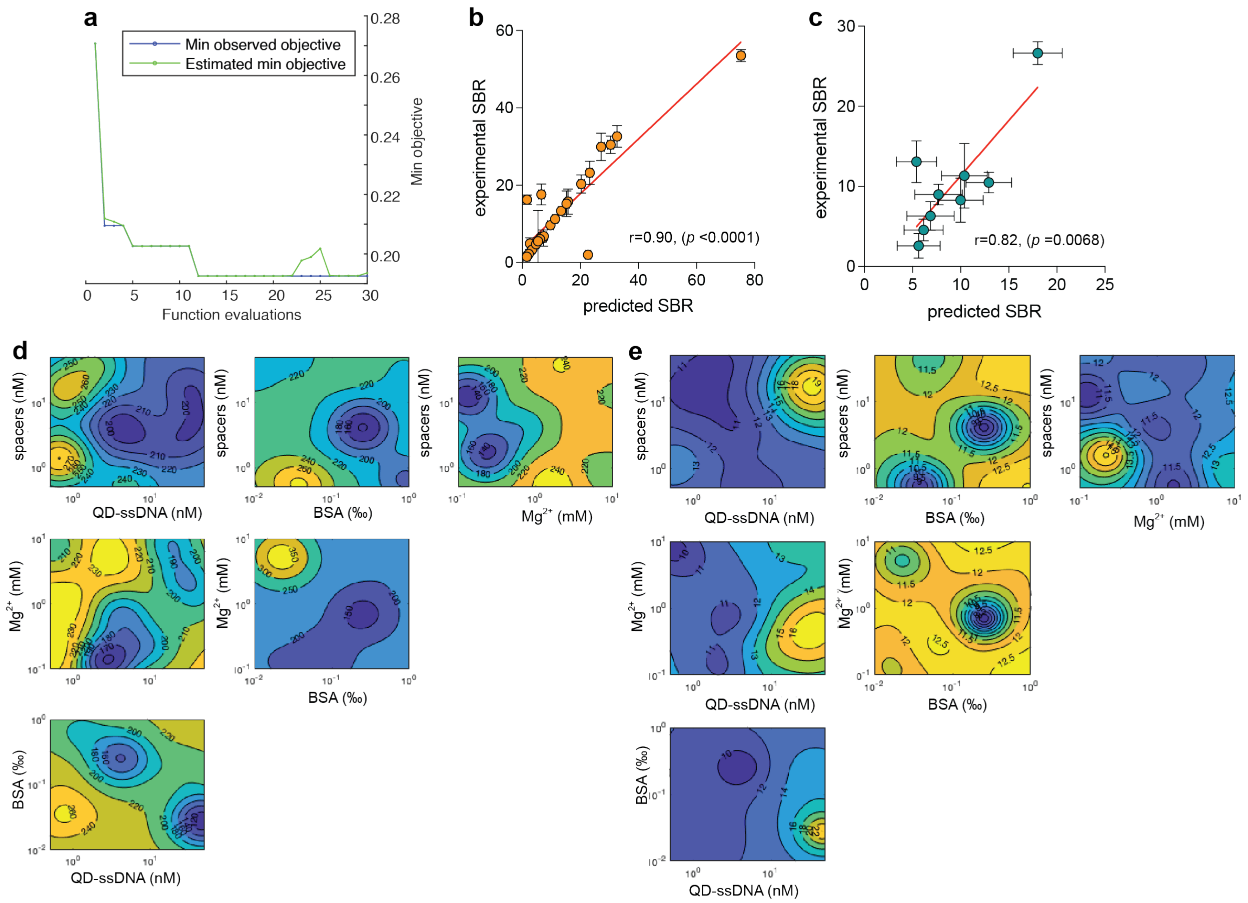


**Figure S19. Validation of model-informed assay optimization. (a)** Objective loss *versus* number of model runs. **(b,c)** Experimental SBR *versus* the SBR predicted by the model is shown (b) for the training group data and (c) test group data. The intraclass correlation coefficient (ICC) for agreement between the predicted results of the test group from the model built by the training group and the experimental value of the test group was good (0.69), reflecting a reliable model. The Pearson r correlation coefficient is indicated in the graph with the *p*-value for the statistical significance of the correlation test. The data points show the mean of technical triplicates, with the red line indicating a linear regression. Y-axis error bars represent the standard deviation of experimental technical triplicates. X-axis error bars represent the 95% confidence interval of the prediction. **(d,e)** Predicted trajectory of (d) signal counts and (e) background counts.

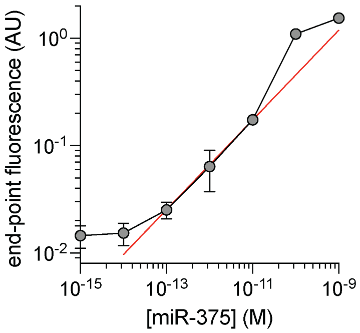


**Figure S20. Quantification of DNFs generated from miR-375 and padlock probes measured using ensemble SYBR Green fluorescence**. Relative fluorescence intensity is shown at each miR-375 concentration. Error bars represent the standard deviation from three technical replicates. The red line represents a linear fit of data points after the log-log transformed data ($R^2$ = 0.90). AU = arbitrary units.

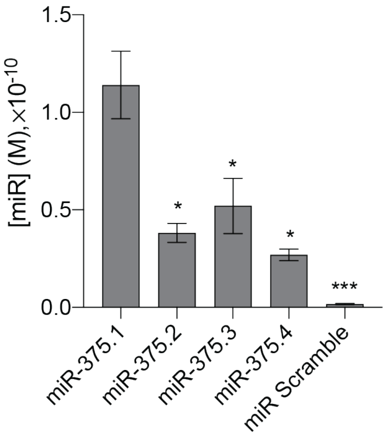


**Figure S21. Characterization of sequence specificity of iQ-Flow for miR-375 isomiRs**. The four prevalent miR-375 isoforms (miR-375.1, miR-375.2, miR-375.3, miR-375.4)[3] were analyzed at 100 pM using iQ-Flow. Detected effective concentrations were calculated using a standard curve generated using the most prevalent isoform, miR-375.1, for which the padlock probe template was designed. Bar height and error bars represent the mean and standard deviation for technical triplicates. These isomiR sequences (**Table S1**) were chosen from next-generation sequencing data of exosomal RNA extracts from plasma, revealing polymorphisms primarily at the 3' end.[3] These findings show that using the padlock DNA probe, we could detect significant differences between the perfect match and its isoform (Student's t-test, *: $p < 0.05$, and *: $p < 0.001$). However, a circular DNA template reported by Smith *et. al.* is unable to differentiate between the miR-375.1 and miR-375.4.[4] This outcome suggests that the padlock method can enhance the specificity of assays for future applications necessitating the precise measurement of a single isoform with high specificity.

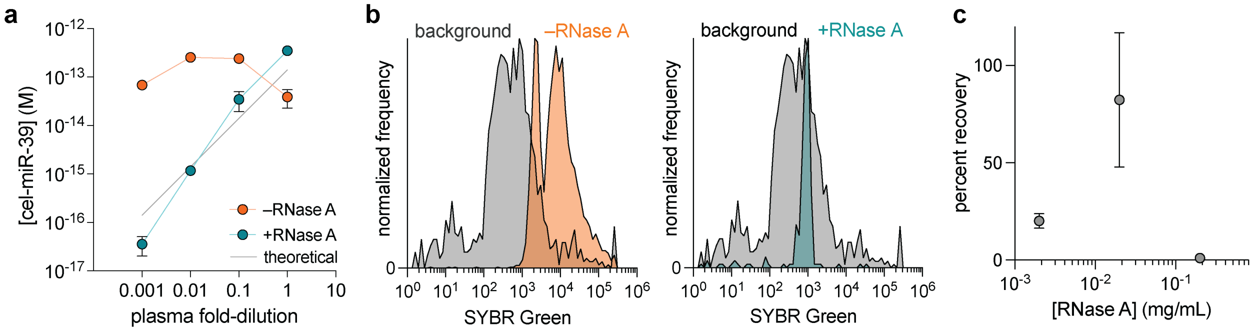


**Figure S22. Impact of digestion of residual RNA in plasma extracts on iQ-Flow assays**. **(a)** iQ-Flow quantification of spike-in cel-miR-39 at different plasma dilutions, showing that RNase A is needed to yield a count proportional to dilution factor slope (slope = 1.02; $R^2$ = 0.88). The gray line represents the theoretical concentration calculated from RT-qPCR. The error bars represent the standard deviation of technical triplicates. **(b)** Histograms of event intensity of SYBR-labeled cel-miR-39-derived DNFs with (green) and without (orange) RNase treatments in plasma extract solution without cel-miR-39 spike-in. The grey DNF histograms represent SYBR intensity in water samples with reaction buffer and no cel-miR-39 spiked in. **(c)** Recovery percentage for spike-in cel-miR-39 at indicated RNase A concentrations. Dot and error bars represent the mean and S.D. for technical triplicates.

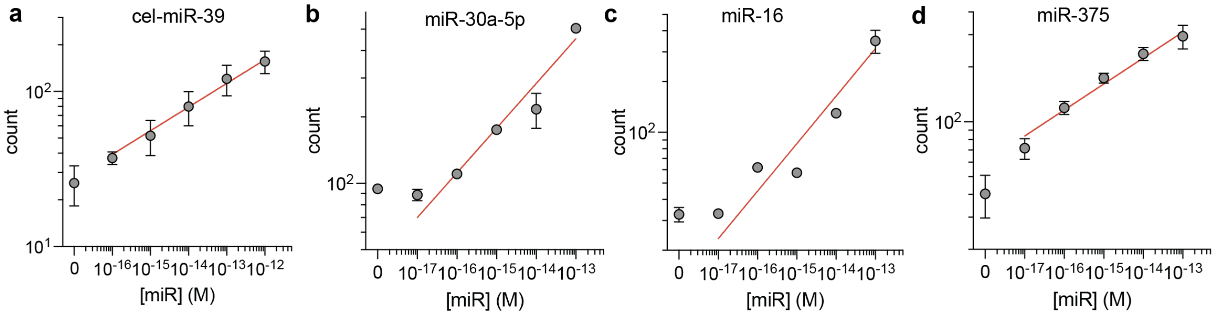


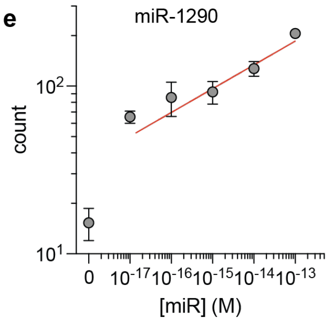


**f**

| miR sequence | Slope | LOD (fM) | $R^2$ |
|---|---|---|---|
| cel-miR-39 | 0.138 | 4.54 | 0.946 |
| miR-30a-5p | 0.121 | 0.257 | 0.982 |
| miR-16 | 0.114 | 0.656 | 0.943 |
| miR-375 | 0.196 | 0.151 | 0.992 |
| miR-1290 | 0.114 | 0.605 | 0.969 |

**Figure S23. Standard curve of iQ-Flow assay using synthetic miRs**. Each point represents mean counts of five biological replicates of **(a)** cel-miR-39, **(b)** miR-30a-5p, **(c)** miR-16, **(d)** miR-375, and **(e)** miR-1290 for concentrations of 0, $10^{-17}$, $10^{-16}$, $10^{-15}$, $10^{-14}$, $10^{-13}$, and $10^{-12}$ (M) with error bars indicating the S.E.M. $N$ = 3. The red line indicates the linear regression of the log-log transformed data. **(f)** The table reports the fitting results, including the slope and LOD of the standard curves.

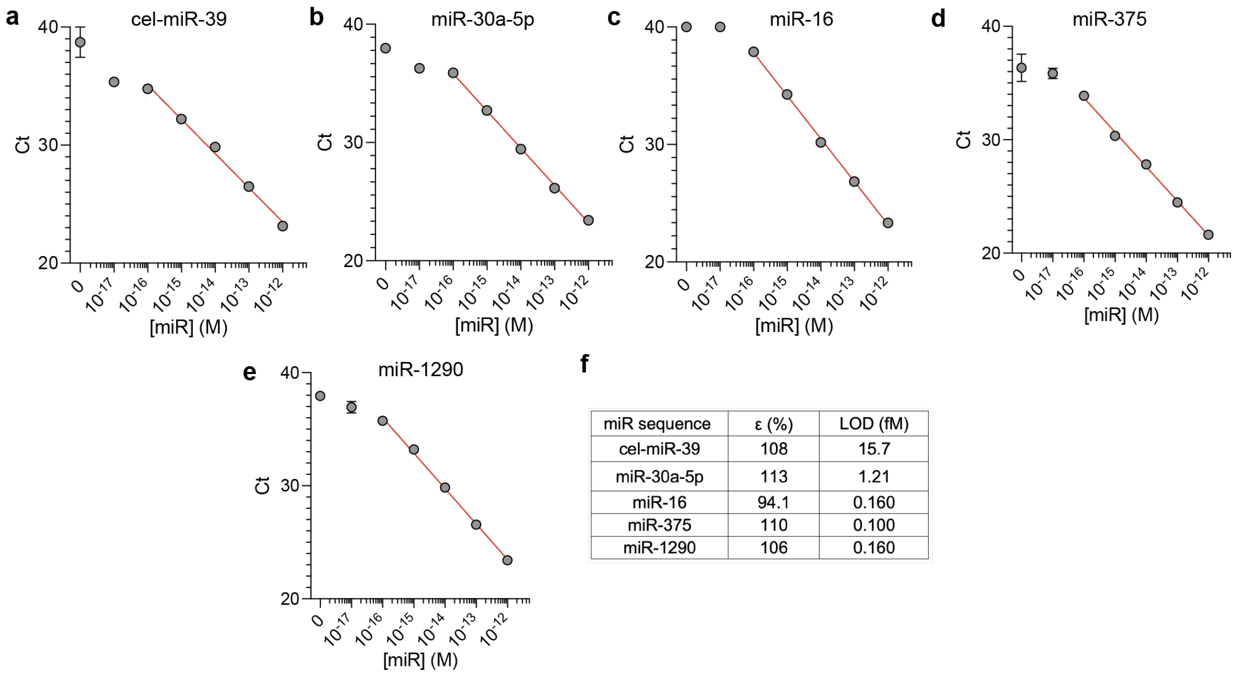


f

| miR sequence | ε (%) | LOD (fM) |
|---|---|---|
| cel-miR-39 | 108 | 15.7 |
| miR-30a-5p | 113 | 1.21 |
| miR-16 | 94.1 | 0.160 |
| miR-375 | 110 | 0.100 |
| miR-1290 | 106 | 0.160 |

**Figure S24. Standard curve of RT-qPCR assay using synthetic miRs**. Each point represents the mean count of biological triplicates of **(a)** cel-miR-39, **(b)** miR-30a-5p, **(c)** miR-16, **(d)** miR-375, and **(e)** miR-1290 for concentrations of 0, $10^{-17}$, $10^{-16}$, $10^{-15}$, $10^{-14}$, $10^{-13}$, and $10^{-12}$ (M) with error bars indicating S.E.M. from biological triplicates. The red line indicates the linear regression from semi-log transformed miR concentration *versus* threshold cycle (Ct). **(f)** The table reports the fitting results, including assay efficiency (ε) and LOD of the standard curves.

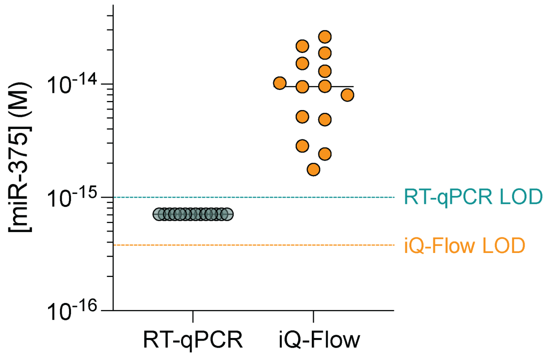


**Figure S25. Measured miR-375 concentration from plasma extracs for RT-qPCR and iQ-Flow**. Each point represents the mean of biological triplicates. The horizontal black line indicates the median value of the quantified miR-375 concentration in the samples. The dashed line (green, RT-qPCR; orange, iQ-Flow) indicates assay LOD (RT-qPCR: 1.00 fM, iQ-Flow: 0.317 fM). RT-qPCR quantified miR-375 level is below the assay LOD. For visualization, values below LOD were substituted with LOD/√2.[5]

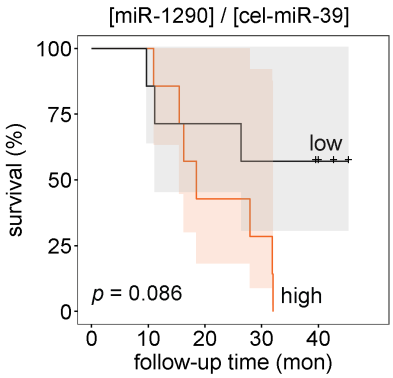


**Figure S26**. **Kaplan-Meier curves show a less survival association with the miR-1290 biomarkers, normalized to cel-miR-39, detected by iQ-Flow**. The orange line and shaded area indicate groups classified as high-risk while the black lines indicate the low-risk group. *N* = 14.

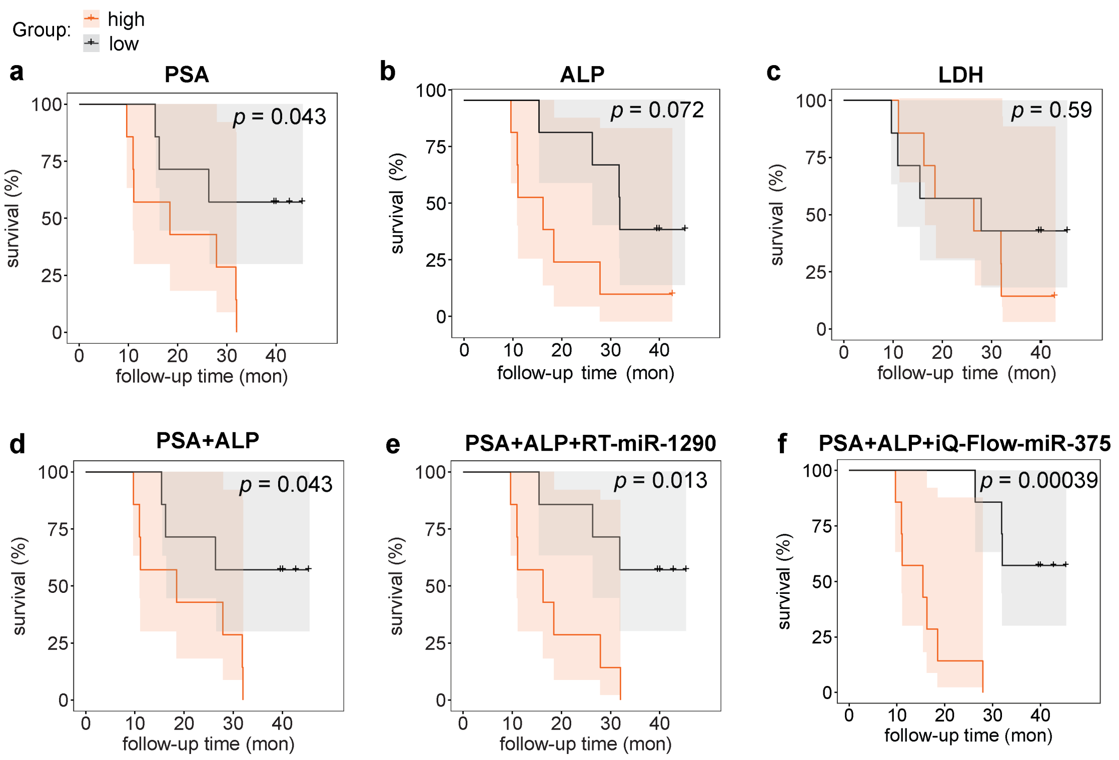


**Figure S27. Kaplan-Meier curves show association between therapy failure time and biomarker levels.** Correlations are shown for **(a)** prostate-specific antigen (PSA), **(b)** alkaline phosphatase (ALP), **(c)** lactate dehydrogenase (LDH), **(d)** PSA+ALP, **(e)** PSA+ALP+normalized miR-1290 quantified by RT-qPCR, and **(f)** PSA+ALP+normalized miR-375 quantified by iQ-Flow. The orange line and shaded area indicate groups classified as high-risk, using a multivariate model, while the black lines indicate the low-risk group. $N = 14$.

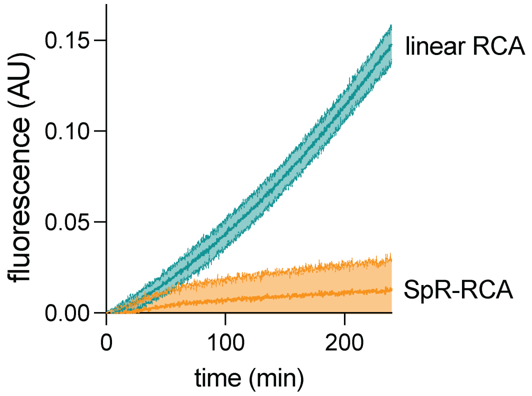


**Figure S28. Reduced RCA non-specificity using padlock probe ligation.** Data show real-time fluorescence intensity during DNF generation with 0 M miR spike-in by RCA using a pre-ligated circular template (green line; linear RCA) or a linear padlock probe circularized using SplintR (SpR) ligation (orange line; SpR-RCA). The shaded area indicates standard deviation from technical triplicates. AU = arbitrary units.

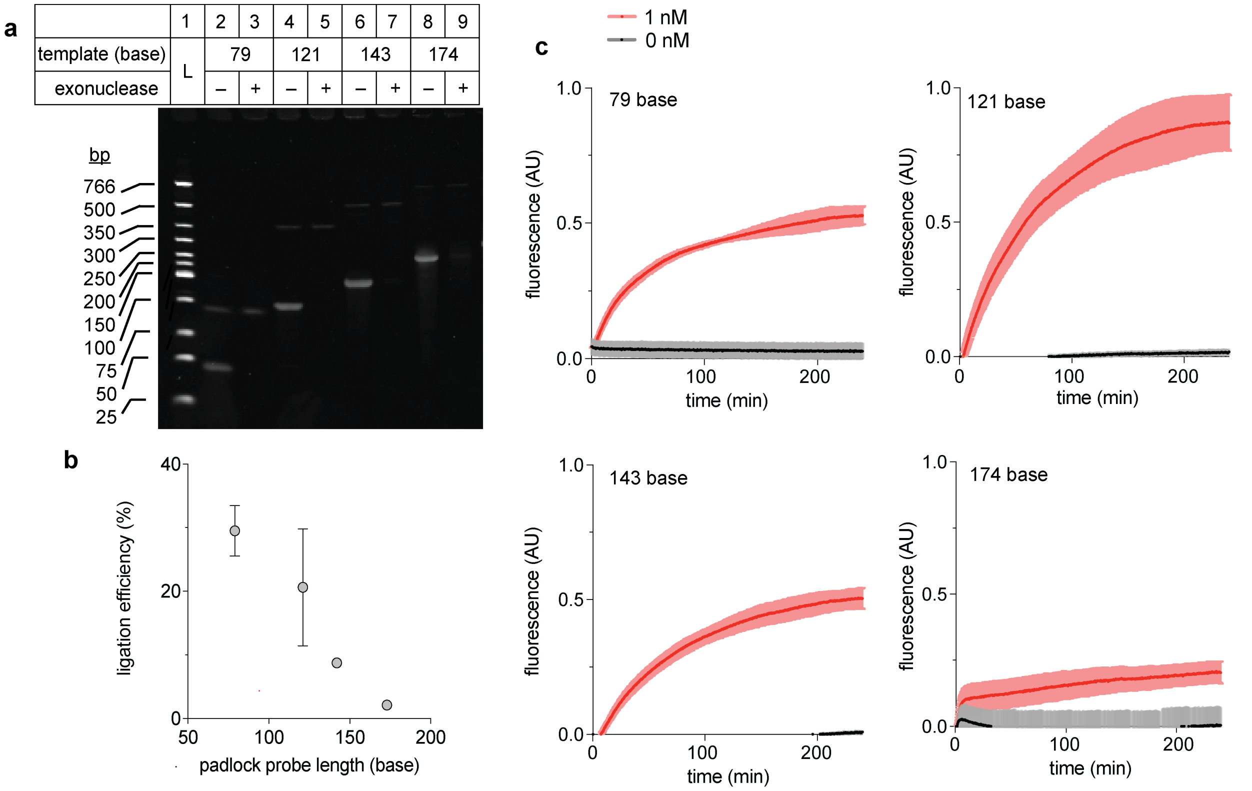


**Figure S29**. **Evaluation of the impact of padlock probe length. (a)** Representative image of 10% polyacrylamide gel after electrophoresis of a low molecular weight DNA ladder (lane 1), circularized template of different lengths (79, 121, 143, and 174 base) without exonuclease I digestion (lanes 2, 4, 6, and 8) or with digestion (lanes 3, 5, 7, and 9). **(b)** Ligation efficiency of different lengths of padlock probes was quantified from the fluorescent intensity of gel electrophoresis. Error bar indicates S.D. *N* = 3. **(c)** Quantitative RCA results of miR-375 at 1 nM (red) or 0 nM (black). The shaded area indicates S.D. *N* = 3. AU = arbitrary units.

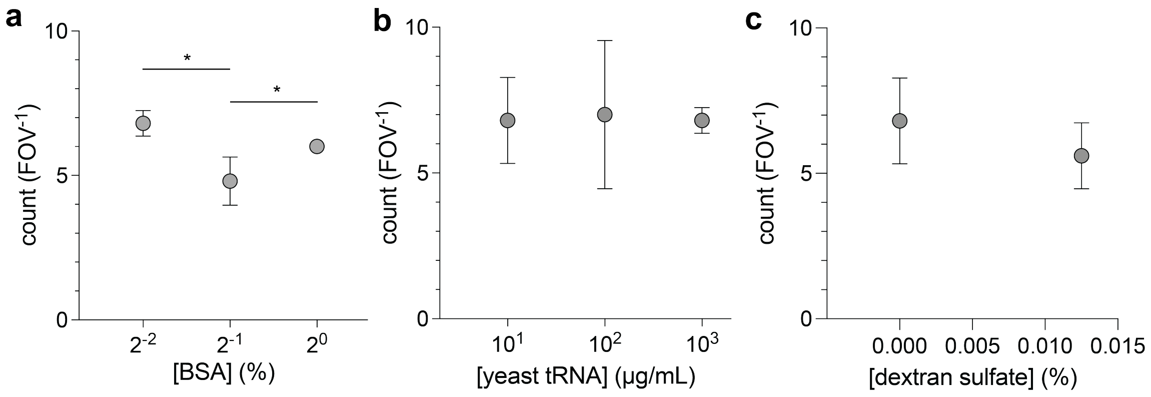


**Figure S30. Optimizing blocking agent cocktails for in-solution assays using 0 nM DNF solution.** Graphs show in-solution assay off-target count dependence on concentrations of **(a)** BSA, **(b)** yeast tRNA and **(c)** dextran sulfate. Error bars indicate S.D. $N = 3$. * indicates $p < 0.05$ by Student's t-test.

### 2. Supporting Tables

#### Table S1. Oligonucleotide sequences

| Name | Sequence |
|---|---|
| miR-375.1 | rUrUrU rGrUrU rCrGrU rUrCrG rGrCrU rCrGrC rGrUrG rA |
| miR-375.2 | rUrUrU rGrUrU rCrGrU rUrCrG rGrCrU rCrGrC rGrUrG |
| miR-375.3 | rUrUrU rGrUrU rCrGrU rUrCrG rGrCrU rCrGrC rGrUrG rU |
| miR-375.4 | rUrUrU rGrUrU rCrGrU rUrCrG rGrCrU rCrGrC rGrU |
| Scramble miR | rUrGrC rUrArA rGrGrU rCrCrG rUrGrU rCrCrA rUrArU rC |
| miR-30a-5p | rUrGrU rArArAr CrArUrCrCrU rCrGrA rCrUrG rGrArA rG |
| miR-16 | rUrArG rCrArG rCrArC rGrUrA rArArU rArUrU rGrGrC rG |
| miR-1290 | rUrGrG rArUrU rUrUrU rGrGrA rUrCrA rGrGrG rA |
| cel-miR-39 | rUrCrA rCrCrG rGrGrU rGrUrA rArArU rCrArG rCrUrU rG |
| miR-375 RCA Template (79 nt) | /5Phos/ CAA CAA CCA ACA AAC ACA GAA TGC TCA CGC GAG CCG AAC GAA CAA ACC TCA GCA ACA CCA AAC AAC AAA C |
| miR-375 RCA Template (121 nt) | /5Phos/ GAA CGA ACA AAC ACA GAT TAA ACG TGC CAT ACC ACA ACT AGA CGA CAC ATA CCG AAT CAC AAC ACC GAG ATG ACT ACC AGA CAA CAA CAG AAC ACC TCA CAT CGA ACC AAT CAC GCG AGC C |
| miR-375 RCA Template (143 nt) | /5Phos/ GAA CGA ACA AAC AGA TTA AAC GTG CCA TAC CAC AAC TAG ACG ACA CAT ACC GAA TCA CAA CAC CGA GAT GAC TAC CAG ACA ACA ACA GAA CAC CTC ACA TCG AAC CAC AAC CAC TAA CCG CAT GCA TAC GAT CAC GCG AGC C |
| miR-375 RCA Template (174 nt) | /5Phos/ GAA CGA ACA AAC AAC AGA TTA AAC GTG CCA TAC CAC AAC TAG ACG ACA CAT ACC GAA TCA CAA CAC CGA GAT GAC TAC CAG ACA ACA ACA GAA CAC CTC ACA TCG AAC CAC AAC CAC TAAC CGC ATG CAT ACG AAC AAC CAG ATT AAA CGT GCC ATA CCA AAT CAC GCG AGC C |
| C1: miR-1290 RCA Template | /5Phos/ AAA AAT CCA ACA AAC CGA GAT GAC TAC CAG ACA ACA ACA CCG AGA TGA CTA CCA GAC AAC AAC ACC GAG ATG ACT ACC AGA CAA CAA CAC CGA GAT GAC TAC CAG ACA AAC TCC CTG ATC C |
| C2: miR-30a-5p RCA Template | /5Phos/GGA TGT TTA CAC ATA GAC GAC ACA TAC CGA ATC ACA ACT AGA CGA CAC ATA CCG AAT CAC AAC TAG ACG ACA CAT ACC GAA TCA CAA CTA GAC GAC ACA TAC CGA ATC AAC TTC CAG TCG A |
| C3: miR-16 RCA Template | /5Phos/ ACG TGC TGC TAC AAG AAC ACC TCA CAT CGA ACC ACA ACA GAA CAC CTC ACA TCG AAC CAC AAC AGA ACA CCT CAC ATC GAA CCA CAA CAG AAC ACC TCA CAT CGA ACC AAC ACC AAT ATT T |
| C4: miR-375 RCA Template | /5Phos/ GAA CGA ACA AAC AAC CGA GAT GAC TAC CAG ACA ACA ACA CCG AGA TGA CTA CCA GAC AAC AAC ACC GAG ATG ACT ACC AGA CAA CAA CAC CGA GAT GAC TAC CAG ACA AAT CAC GCG AGC C |
| C5: cel-miR-39 RCA Template | /5Phos/ CAC CCG GTG ACA CAC TAA CCG AAG GCA TAC GAA CAA CCA CTA ACC GAA GGC ATA CGA ACA ACC ACT AAC CGA AGG CAT ACG AAC AAC CAC TAA CCG AAG GCA TAC GAA ACA AGC TGA TTT A |

**Table S1 (continued)**

| | |
|---|---|
| iQ-Flow miR-30a-5p RCA Template | /5Phos/ GGA TGT TTA CAC ACA GAT TAA ACG TGC CAT ACC ACA ACT AGA CGA CAC ATA CCG AAT CAC AAC ACC GAG ATG ACT ACC AGA CAA CAA CAG AAC ACC TCA CAT CGA ACC AAC TTC CAG TCG A |
| iQ-Flow miR-16 RCA Template | /5Phos/ ACG TGC TGC TAC ACA GAT TAA ACG TGC CAT ACC ACA ACT AGA CGA CAC ATA CCG AAT CAC AAC ACC GAG ATG ACT ACC AGA CAA CAA CAG AAC ACC TCA CAT CGA ACC AAC ACC AAT ATT T |
| iQ-Flow miR-1290 RCA Template | /5Phos/ AAA AAT CCA ACA ACA GAT TAA ACG TGC CAT ACC ACA ACT AGA CGA CAC ATA CCG AAT CAC AAC ACC GAG ATG ACT ACC AGA CAA CAA CAG AAC ACC TCA CAT CGA ACC AAC TCC CTG ATC C |
| iQ-Flow cel-miR-39 RCA Template | /5Phos/ CAC CCG GTG ACA CAG ATT AAA CGT GCC ATA CCA CAA CTA GAC GAC ACA TAC CGA ATC ACA ACA CCG AGA TGA CTA CCA GAC AAC AAC AGA ACA CCT CAC ATC GAA CCA ACA AGC TGA TTT A |
| DBCO-L1 | /5DBCOTEG/ CAG ATT AAA CGT GCC ATA CC |
| DBCO-R1' | /5DBCOTEG/ CCG TAA CGA GCG TCC CTT GC |
| DBCO-S1 | /5DBCOTEG/ CCG GAT AAC TTC AGT AAA CC |
| DBCO-R2' | /5DBCOTEG/ CGA ACT CAG ACT CAC CCT AC |
| DBCO-R3' | /5DBCOTEG/ CGG AGC GTA GCG GAA TCT GC |
| DBCO-R4' | /5DBCOTEG/ TAC TCA CAC CGA GAC GAC AC |
| DBCO-R5' | /5DBCOTEG/ CCA CTA CCG AAA GTT CCG AA |
| Alexa546-L1 | /5Alex546N/ CAG ATT AAA CGT GCC ATA CC |
| Biotin-L3 | /5BiotinTEG/ ACC GAG ATG ACT ACC AGA CA |
| Spacer L1–R1 | CAG ATT AAA CGT GCC ATA CCT ATA TAC ATT CTC TAA ATA CTA TAC ATT GAA TCT ATT AAC AAA GTT ATT AAT TTA ATT ATG CAA GGG ACG CTC GTT ACG G |
| Spacer L2–R2 | TAG ACG ACA CAT ACC GAA TCA CGA TTT ATC CTC CTT ATT AAT CTA ATA TTC ATT TAT AAC CAT AAA ATC GTT TAT TTA TCG TAG GGT GAG TCT GAG TTC G |
| Spacer L3–R3 | ACC GAG ATG ACT ACC AGA CAA CAA TGA TAA TTT ATT TTT AAT CTA ATT TTC TTT TAT AAC CAT AAA ATC GTT TAT TTA TCG CAG ATT CCG CTA CGC TCC G |
| Spacer L4–R4 | AGA ACA CCT CAC ATC GAA CCA CGA TTT ATC CTC CTT ATT AAT CTA ATA TTC ATT TAT AAC CAT AAA ATC GTT TAT TTA TCT CGG AAG CGT GTA TAG TGG C |
| Spacer L5–R5 | CAC TAA CCG CAT GCA TAC GAA CGA TTT ATC CTC CTT ATT AAT CTA ATA TTC ATT TAT AAC CAT AAA ATC GTT TAT TTA TCT TCG GAA CTT TCG GTA GTG G |
| L1 | CAG ATT AAA CGT GCC ATA CC |
| L2 | TAG ACG ACA CAT ACC GAA TC |
| L3 | ACC GAG ATG ACT ACC AGA CA |
| L4 | AGA ACA CCT CAC ATC GAA CC |
| L5 | CAC TAA CCG CAT GCA TAC GA |

/5Phos/ indicates 5’ phosphate group modification.
/5DBCOTEG/ indicates 5’-dibenzocyclooctyne (DBCO) modification with TEG spacer.
/5Alex546N/ indicates 5’ modifications with the dyes Alexa Fluor 546.
/5BiotinTEG/ indicates 5’-biotin modifications with TEG spacer.

**Table S2. Mean spectral characteristics of DNFs labeled with single-color QDs.**

| QD color | $\lambda_{peak}$ (nm) | Full-width at half-maximum (FWHM, nm) |
|---|---|---|
| 445 | 449.1 | 42.0 |
| 525 | 519.9 | 38.8 |
| 570 | 566.8 | 36.1 |
| 605 | 595.9 | 31.7 |
| 625 | 622.1 | 39.7 |

**Table S3. Kaplan-Meier significance metrics from iQ-Flow miR panels using indicated normalization sequences.**

| normalization sequence | miR-375 (*p*-value) | miR-1290 (*p*-value) | miR-375 + miR-1290 (*p*-value) |
|---|---|---|---|
| cel-miR-39 | 0.0011 | 0.086 | 0.0011 |
| miR-30a-5p | 0.098 | 0.098 | 0.098 |
| miR-16 | 0.060 | 0.30 | 0.060 |
| cel-miR-39, miR-16 | 0.060 | 0.30 | 0.0048 |
| cel-miR-39, miR-30a-5p | 0.0011 | 0.11 | 0.0011 |
| miR-16, miR-30a-5p | 0.060 | 0.17 | 0.17 |
| cel-miR-39, miR-30a-5p, miR-16 | 0.0048 | 0.30 | 0.0048 |

**Table S4. Kaplan-Meier significance metrics from RT-qPCR miR panels using indicated normalization sequences.**

| normalization sequence | miR-1290 (*p*-value) |
|---|---|
| cel-miR-39 | 0.060 |
| miR-30a-5p | 0.22 |
| miR-16 | 0.53 |
| cel-miR-39, miR-16 | 0.62 |
| cel-miR-39, miR-30a-5p | 0.33 |
| miR-16, miR-30a-5p | 0.33 |
| cel-miR-39, miR-30a-5p, miR-16 | 0.62 |

**Table S5. Univariate and multivariate hazard ratio comparison**

| variables | hazard Ratio | *p* |
|---|---|---|
| PSA | 2.84 (0.96 – 8.35) | 0.02 |
| LDH | 1.05 (0.60 – 1.84) | 0.9 |
| ALP | 2.46 (1.22 – 4.93) | 0.01 |
| iQ-Flow-miR-375 | 2.33 (1.05 – 5.13) | 0.03 |
| PSA+ALP+iQ-Flow-miR-375 | 2.72 (1.32 – 5.51) | 0.004 |

### 3. Supporting Materials and Methods

**Materials.** Unless stated otherwise, reagents were obtained from Sigma-Aldrich and used without further purification. Oleylamine (OLA, > 80%), 1-octadecene (ODE, 90%), and oleic acid (OAc, 90%) were from Acros Organics. Zinc acetate ($Zn(Ac)_2$, 99.9%) was from Alfa Aesar. DNA and RNA oligonucleotides with sequences shown in **Table S1** were from Integrated DNA Technologies. CircLigase II was from Lucigen. Deoxynucleotide Solution Mix (dNTP), Φ29 DNA polymerase, Murine RNase Inhibitor, SplintR Ligase, Monarch RNase A, *E. coli* Exonuclease I, Low Molecular Weight DNA Ladder, and nuclease-free water were from New England Biolabs. TaqMan miR assays, SUPERase-In RNase Inhibitor, SYBR Green I Nucleic Acid Gel Stain (SYBR Green), SYBR Gold Nucleic Acid Gel Stain (SYBR Gold), BSA, sodium acetate, glacial acetic acid (>99.7%), 6× DNA Gel Loading Dye, TRIzol LS reagent, glycogen, glycerol, and 8-well Lab-Tek chambered covered glass were from ThermoFisher Scientific. Falcon round-bottom polystyrene test tubes were from Fisher Scientific. TBE buffer (10×) and 10% polyacrylamide TBE-urea gels were from Bio-Rad Laboratories. Ethanol was from Decon Labs. PBS was from Corning. Monomethoxy monosuccinimidyl ester poly(ethylene glycol) (mPEG5000-NHS, 5 kDa) and monobiotin monosuccinimidyl ester poly(ethylene glycol) (biotin-PEG5000-NHS, 5 kDa) were from Nanocs, Inc. Flow-Check Fluorospheres were from Beckman Coulter. Citrate vacutainer tubes were from BD. 50-well chambered coverglass was from Electron Microscopy Sciences. Ultrathin carbon film TEM grid, and a diced silicon wafer for SEM imaging were from Ted Pella, Inc. Streptavidin was from ProSpec-Tany TechnoGene Ltd. ExoQuick kit was from System Biosciences. QD685 (8.5 nm) and QD685 (3.3 nm) in **Figure 4c-4f** were used from the same batch used in our previous manuscript.[6]

**Precursor Synthesis for QD synthesis.** Cadmium behenate ($Cd(BAc)_2$) was prepared as follows: BAc (10 mmol) was dissolved in methanol (200 mL) with the addition of tetramethylammonium hydroxide (TMAH) solution (~3 mL), followed by sonication and stirring for 15 min until complete dissolution. The solution was centrifuged, and the colorless supernatant was transferred to a beaker and stirred at 60 °C until the TMAH odor dissipated. In a separate beaker, cadmium chloride (5 mmol) was dissolved in methanol/water (20 mL, 4:1 v/v) and added dropwise to the BAc solution under vigorous stirring. The mixture was stirred for an additional hour, and the resulting white $Cd(BAc)_2$ precipitate was collected by vacuum filtration, washed repeatedly with methanol, and dried overnight on the funnel under ambient air. 0.2 M cadmium oleate solution: cadmium oxide (CdO, 10 mmol), OAc (40 mmol), and ODE (29.45 mL) were added to a 100 mL three-neck round-bottom flask, degassed under vacuum at

100 °C to remove volatiles, filled with $N_2$, heated to 240 °C to dissolve the Cd precursor, and then cooled to room temperature. OLA (6.58 mL, 20 mmol) was subsequently added, and the mixture was degassed under vacuum at 100 °C for 30 min, backfilled with $N_2$, and cooled to room temperature. For Cd and Zn precursors for shell growth, $Cd(Ac)_2$ or $Zn(Ac)_2$ (1 mmol) was dissolved in OLA (10 ml) at 100 °C. For S precursor for shell growth, S powder (1 mmol) was dissolved in ODE (10 ml) at 150 °C.

**QD525, QD570, QD605, and QD625 synthesis.** QDs were synthesized with a core/shell CdSe/CdZnS structure by similar methods modified from previous reports.[7,8] QDs with emission at 525, 570, 605, and 625 nm were synthesized with CdSe core sizes of 2.2, 2.2, 3.2, and 3.7 nm, respectively, using a heat-up reaction between $Cd(BAc)_2$ (1 mmol), selenium dioxide ($SeO_2$, 1 mmol), and 1,2-hexadecanediol (HDD, 1 mmol) in ODE (20 mL). After purification, a shell of CdZnS was grown on these QDs epitaxially through layer-by-layer shell growth with 0.8-monolayer (ML) increments by dropwise addition of an S precursor, followed by the Cd/Zn precursor ($Cd(Ac)_2$ or $Zn(Ac)_2$). The growth of the first S layer was initiated at 120 °C. After 10 min, the Cd precursor was added and allowed to react for 10 min. The solution was then heated to 180 °C and the Zn precursor was added and allowed to react for another 10 min. The reaction temperature was raised in 10 °C increments between each precursor addition until reaching a maximum of 200 °C. The shell composition for QD525 comprised 0.8 ML $Cd_{0.5}Zn_{0.5}S$, 1.2 ML $Cd_{0.2}Zn_{0.8}S$, and 0.2 ML ZnS. For QD570, the shell was composed of 2.4 ML CdS, 0.8 ML $Cd_{0.7}Zn_{0.3}S$, and 0.8 ML ZnS. For QD605 and QD625, the shell composition comprised 2.4 ML CdS, 0.8 ML $Cd_{0.8}Zn_{0.2}S$, and 1.5 ML ZnS. After the reaction, QDs were purified by precipitation with methanol and acetone three times and finally dispersed in hexane. TEM-determined diameters after shell growth were, respectively, 3.3 nm, 4.3 nm, 5.7 nm, and 6.7 nm.

**QD445 synthesis.** These QDs were prepared from ZnSe cores that were cation exchanged with cadmium to generate $Cd_xZn_{1-x}Se$ nanocrystals**.** To synthesize ZnSe, a flask containing OLA (20 mL) was dried under vacuum at 100 °C for 30 min and then charged with nitrogen. The solution was heated to 300 °C before slow addition of a trioctylphosphine selenide solution (0.5 mL, 1 M) diluted in tri-*n*-octylphosphine (TOP, 0.8 mL). A zinc solution was prepared by diluting diethylzinc in hexane (0.5 mL,1 M) in TOP (0.8 mL) before rapid injection into the selenium solution. The temperature was maintained at 290 °C, and aliquots were collected to monitor growth until the size reached 3.2 nm. The reaction mixture was cooled to room temperature, and the crude solution (1.5 mL) was mixed with ethanol (20 mL) and isopropanol (20 mL) and centrifuged at 7000 *g* for 4 min. The precipitate was dissolved in hexane (1.5 mL) and the ethanol/isopropanol precipitation was repeated two more times before the products were finally dispersed in hexane. The purified ZnSe QDs (120 nmol) were then dispersed in a mixture of ODE (3 mL), OLA (5 mL), TOP (3 mL), and hexylphosphonic acid (22 mg) and heated to 220 °C. A solution of cadmium oleate (0.2 M, 44 μL) was then injected to yield $Cd_xZn_{1-x}Se$ QDs. After 15 min, the reaction solution was cooled to room temperature. The QDs were purified by precipitation with methanol and acetone before dispersion of the precipitate in hexanes, a process that was repeated three times. For shell growth, $Cd_xZn_{1-x}Se$ QDs (80 nmol) were dissolved in ODE (4 mL) and OLA (2 mL) and heated to 120 °C. Shell growth was performed as described above to generate 2.4 ML $Cd_{0.2}Zn_{0.8}S$ and 2.0 ML of ZnS.

**QD700 synthesis.** These QDs were synthesized from $CdSe_xS_{1-x}$ cores that were cation exchanged with mercury to generate $Hg_xCd_{1-x}Se_yS_{1-y}$ QDs following our previously published protocol.[9] First, $CdSe_xS_{1-x}$ cores were synthesized by heating a vacuum-dried mixture (100 °C, 1 h) of $Cd(BAc)_2$ (0.2 mmol), $SeO_2$ (0.13 mmol), S powder (0.07 mmol), and HDD (0.2 mmol) in ODE (4 mL) to 230 °C under nitrogen, allowing 15 min reaction at this temperature. The resulting cores had a diameter of 2.8 nm with the first absorption peak at 506 nm. The QDs were purified and dispersed in chloroform (100 nmol, 5 mL) before the addition of a mercury acetate solution in OLA (0.2 M) with a 2-fold excess of mercury relative to the total amount of cadmium in the QD. After~5 min, cation exchange was quenched by injection of 1-octanethiol (100 μL), and the QDs were precipitated with a methanol/acetone mixture (1:1 v/v; 20 mL). The QDs were further purified by three cycles of dispersion in hexane (10 mL) with OLA (100 μL) and OLAc (100 μL), followed by precipitation with methanol/acetone (20 mL). The resulting $Hg_xCd_{1-x}Se_yS_{1-y}$ QDs were stored as a stock solution in hexane. CdZnS shells using the methods described above QDs to generate 2.4 ML CdS, 0.8 ML $Cd_{0.8}Zn_{0.2}S$, and 0.8 ML ZnS. TEM-determined diameters after shell growth were 4.5 nm.

**Polymer coating of QDs.** QDs were coated through the reported protocol.[8] Purified QDs in hexane were phase-transferred to *N*-methylformamide (NMF) by dropwise addition of TMAH (25 wt % in methanol) at 100 equivalents to QD surface atoms. A multidentate polymer, polyacrylamide(histamine-*co*-triethylene glycol-*co*-azidotriethylene glycol) (P-IM-$N_3$), dissolved in anhydrous dimethyl sulfoxide (10.0 mg/mL, 200 μL) was mixed with the hydroxide-coated QDs in NMF (4.0 μM, 250 μL) at a 5:1 ratio of imidazoles to QD surface atoms. Under a nitrogen atmosphere using a Schlenck line, the solution was stirred at 110 °C for 2 h before precipitation with a mixture of ether and chloroform. The QDs were collected by centrifugation, dried, and dispersed in sodium borate buffer (50 mM; pH 8.5). Excess polymer was removed by centrifugal filtration (Amicon Ultra 50 kDa MWCO) with dilution and filtration repeated six times.

**QD characterization using GPC.** GPC was performed using a GE Healthcare ÄKTApurifier UPC10 with a Cytiva Superose 6 increase 10/300GL column with PBS as mobile phase at a flow rate of 0.5 mL/min. A 100 µL sample loop was used with complete filling for each sample. Product elution was measured by optical density at 254 nm wavelength. QDs dispersed in PBS were centrifuged at 7000 *g* for 5 min and filtered through a 0.2 µm syringe filter before loading into the column. Calibration of elution time was determined using a gel filtration marker kit for protein molecular weights of 29–700 kDa. Time-based absorption data were collected with UNICORN 5.31 Workstation software. Using Origin software (Origin Lab), a Gaussian mixture model was fit to each chromatogram to calculate hydrodynamic diameter values from the peak times using the linear regression of the size calibration of peak elution times.

**Polyacrylamide gel electrophoresis.** Samples were prepared in 1x TBE buffer and loaded into 10% polyacrylamide TBE-urea gels in a Bio-Rad Mini-PROTEAN Tetra Vertical Electrophoresis Cell with 1× TBE buffer. Electrophoresis was performed at 25 V for 1 h, followed by 50 V for 2.5 h. Gels were then incubated for 1 h at room temperature in a 1 : 10000 dilution of SYBR Gold in 1× TBE buffer. Gels were then washed with deionized water and imaged using a Bio-Rad Gel Doc XR+ System with ultraviolet illumination.

**Polyacrylamide–Agarose Gel Electrophoresis.** Horizontal gel electrophoresis using a gel mixture of polyacrylamide and agarose was prepared and run using our published protocols.[9,10]

**Circular template synthesis.** The miR-375 RCA Template (121 nt, 500 nM, **Table S1**) with 5'-phosphoryl modification was circularized using CircLigase II (10 U µL$^{-1}$) in 0.33 M Tris-acetate, 0.66 M potassium acetate, 2.5 mM manganese(II) chloride, 1 M betaine, and 5 mM dithiothreitol at pH 7.5 for 2 h at 60 °C. Unreacted linear DNA was degraded by reaction with Exonuclease I for 2 h at 37 °C before a 10 min incubation at 80 °C, followed by storage in –20 °C until further use. The concentration of the circularized template was quantified using polyacrylamide gel electrophoresis by calibrating fluorescent intensity to concentration**.**

**Linear-RCA reaction.** The circularized miR-375 RCA Template (1nM) (**Table S1**) was mixed with synthetic miR-375 (**Table S1**) at different concentrations at room temperature in a 20 µL solution containing 0.4 U µL$^{-1}$ Φ29 DNA polymerase, 0.5 U µL$^{-1}$ SUPERase-In RNase Inhibitor, 0.5 U µL$^{-1}$ Murine RNase Inhibitor, 1 mM dNTPs, 0.1 mg mL$^{-1}$ BSA, and 0.001% SYBR Gold at pH 7.5). Reactions were performed using a ThermoFisher QuantStudio 3 Real-Time PCR with SYBR Gold fluorescence monitored using LED excitation at 470 nm and emission at 520 ± 10 nm at 15 s intervals. Reactions were allowed to proceed for 4 h at 37 °C prior to polymerase heat inactivation at 80 °C for 10 min.

**Model-informed assay optimization**. The optimization workflow is similar to the reported literature[11,12] and illustrated in **Figure S17**. Following monotonic analysis, we applied the evolutionary operation to search for the most space-filling training group (N=25) across four parameters by iteratively mutating previous results to attain the optimal solution. Following the acquisition of experimental results based on the LHD-suggested set, we further used GPR to help us predict the surrogate model. The hyperparameters of the Matern32 kernel are automatically optimized during the regression by minimizing the five-fold cross-validation loss. (**Figure S19a**). To validate the model, we are then using an additional testing group (N=9) generated again by the evolutionary operation for evaluating the prediction accuracy of the regression model using an agreement test. All the simulation was performed in custom MATLAB code (Mathworks).
